# Functional SARS-CoV-2-specific immune memory persists after mild COVID-19

**DOI:** 10.1101/2020.08.11.20171843

**Authors:** Lauren B. Rodda, Jason Netland, Laila Shehata, Kurt B. Pruner, Peter A. Morawski, Chris Thouvenel, Kennidy K. Takehara, Julie Eggenberger, Emily A. Hemann, Hayley R. Waterman, Mitchell L. Fahning, Yu Chen, Jennifer Rathe, Caleb Stokes, Samuel Wrenn, Brooke Fiala, Lauren Carter, Jessica A. Hamerman, Neil P. King, Michael Gale, Daniel J. Campbell, David Rawlings, Marion Pepper

## Abstract

The recently emerged SARS-CoV-2 virus is currently causing a global pandemic and cases continue to rise. The majority of infected individuals experience mildly symptomatic coronavirus disease 2019 (COVID-19), but it is unknown whether this can induce persistent immune memory that might contribute to herd immunity. Thus, we performed a longitudinal assessment of individuals recovered from mildly symptomatic COVID-19 to determine if they develop and sustain immunological memory against the virus. We found that recovered individuals developed SARS-CoV-2-specific IgG antibody and neutralizing plasma, as well as virus-specific memory B and T cells that not only persisted, but in some cases increased numerically over three months following symptom onset. Furthermore, the SARS-CoV-2-specific memory lymphocytes exhibited characteristics associated with potent antiviral immunity: memory T cells secreted IFN-γ and expanded upon antigen re-encounter, while memory B cells expressed receptors capable of neutralizing virus when expressed as antibodies. These findings demonstrate that mild COVID-19 elicits memory lymphocytes that persist and display functional hallmarks associated with antiviral protective immunity.

## Main Text

The rapid spread of the SARS-CoV-2 beta coronavirus has infected 19 million and killed over 700,000 people worldwide as of early August 2020. Infection causes the disease COVID-19, which ranges in presentation from asymptomatic to fatal. However, the vast majority of infected individuals experience mild symptoms that do not require hospitalization^1^. It is critically important to understand if SARS-CoV-2–infected individuals who recover from mild disease develop immune memory that protects them from subsequent SARS-CoV-2 infections, thereby reducing transmission and promoting herd immunity.

Immunological memory is predominantly mediated by cells of the adaptive immune system. In response to most acute viral infections, B and T cells that can bind viral antigens through their antigen receptors become activated, expand, differentiate and begin secreting effector molecules to help control the infection. Upon resolution of infection, approximately 90% of these virus-specific “effector cells” die, while 10% persist as long-lived “memory” cells^2^. Immune memory cells can produce a continuous supply of effector molecules, as seen with long-lived antibody-secreting plasma cells (LLPCs). In most cases, however, quiescent memory lymphocytes are strategically positioned to rapidly reactivate in response to re-infection and execute effector programs imprinted upon them during the primary response. Upon re-infection, pathogen-specific memory B cells (MBCs) that express receptors associated with antigen experience and the transcription factor T-bet rapidly proliferate and differentiate into IgG^+^ antibody-secreting plasmablasts (PBs)^3–5^. Reactivated T-bet-expressing memory CD4^+^ T cells proliferate, “help” activate MBCs and secrete cytokines (including IFNγ) to activate innate cells^2^. Meanwhile, memory CD8^+^ T cells can kill virus-infected cells directly through the delivery of cytolytic molecules^6^. These quantitatively and qualitatively enhanced virus-specific memory populations coordinate to quickly clear the virus, thereby preventing disease and reducing the chance of transmission.

To infect cells and propagate, SARS-CoV-2 relies on the interaction between the receptor binding domain (RBD) of its spike protein (S) and angiotensin converting enzyme 2 (ACE2) on host cells^7^. Multiple studies have shown that the majority of SARS-CoV-2 infected individuals produce S- and RBD-specific antibodies during the primary response, and RBD-specific monoclonal antibodies can neutralize the virus *in vitro* and *in vivo*^8–10^. Therefore, RBD-specific antibodies would likely contribute to protection against re-infection if expressed by LLPCs or MBCs.

To determine if the above hallmarks of immune protection from viral infection both form and persist in individuals that have experienced mild COVID-19, we assessed their SARS-CoV-2-specific immune responses at one and three months post-symptom onset. Herein we demonstrate that a multipotent SARS-CoV-2-specific immune memory response forms and is maintained in recovered individuals at least for the duration of our study. Furthermore, memory lymphocytes display hallmarks of protective antiviral immunity.

### Return to immune homeostasis after mildly symptomatic COVID-19

To determine if immune memory cells form after mildly symptomatic COVID-19, we collected plasma and peripheral blood mononuclear cells (PBMCs) from 15 individuals recovered from COVID-19 (CoV2^+^) (UW IRB 00009810). The CoV2^+^ group had a median age of 47 and reported mild symptoms lasting a median of 13 days (**E.D. Table 1**). The first blood sample (Visit 1) was drawn at least 20 days after a positive PCR test for SARS-CoV-2 and a median of 35.5 days post-symptom onset. We expect the primary response to be contracting and early memory populations to be generated at this time point, as viral load is cleared approximately 8 days post symptom onset^11^. Participants returned for a second blood draw (Visit 2) a median of 86 days post-symptom onset so we could assess the quantity and quality of the long-lived memory populations (**Fig. 1a**). We compared these samples to samples collected at two time points representing a similar sampling interval in a group of 17 healthy controls (HCs). All HCs were considered to have no prior SARS-CoV-2 infection based on having no detectable plasma SARS-CoV-2 RBD- or S-specific antibodies above three standard deviations (SDs) of the mean of historical negative (HN) plasma samples (**E.D. Fig. 1**). We also included HN PBMC samples that were collected prior to the first human SARS-CoV-2 infection (2016-2019). We included these to control for the possibility that individuals in the HC group had been infected with SARS-CoV-2 (9/17 described having some symptoms associated with SARS-CoV-2 infection) despite their lack of detectable RBD-specific antibodies.

**Figure 1:**
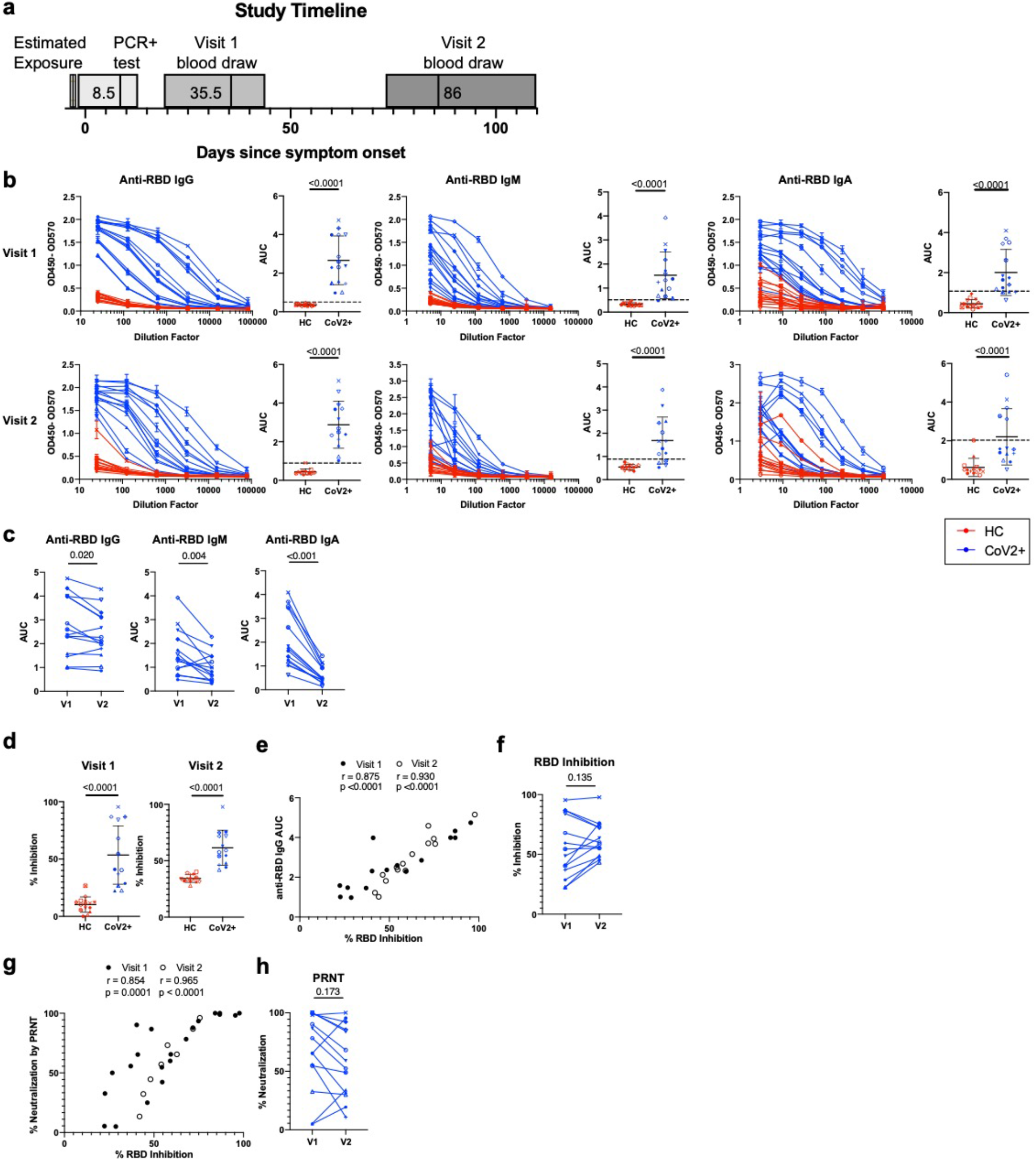
SARS-CoV-2-specific plasma antibodies at two memory time points. **a)** Study timeline. Range indicated by box and median indicated by line for each event. **b)** ELISA dilution curves and AUC for anti-RBD IgG (left), IgM (center), and IgA (right) from healthy control (HC) and previously SARS-CoV-2-infected (CoV2^+^) plasma samples at Visit 1 (V1) and Visit 2 (V2). Dashed line indicates mean + 3 SD of the HC AUC values. Each symbol is a different individual and is consistent throughout the figure. **c)** V2 CoV2^+^ AUC values were normalized to V1 samples run with V2 samples and AUC for each CoV2^+^ individual from V1 and V2 are paired. **d)** Percent inhibition of RBD binding to ACE2 by plasma at 1:10 dilution. **e)** Spearman correlation between percent RBD inhibition at a 1:10 plasma dilution and anti-RBD IgG AUC. **f)** CoV2^+^ percent RBD inhibition at 1:10 plasma dilution normalized and paired as in c). **g)** Spearman correlation between percent RBD inhibition at a 1:10 plasma dilution and percent virus neutralization by PRNT at a 1:80 plasma dilution. **h)** CoV2^+^ percent virus neutralization by PRNT at a 1:80 plasma dilution normalized and paired as in c). Statistical significance for unpaired data determined by two-tailed Mann-Whitney tests and, for paired data, by two-tailed Wilcoxon signed-rank tests. Error bars represent mean and SD (V1 HC n=15, V2 HC n=14, V1 CoV2^+^ n=15, V2 CoV2^+^ n=14).

Populations of activated innate and adaptive immune cells expand in the blood during the primary response to SARS-CoV-2 infection^12^. When an acute viral infection is cleared, the majority of these highly inflammatory cells either die or become quiescent memory cells such that the proportions and phenotypes of total immune cells are indistinguishable from those seen in pre-infection blood samples. Consistent with resolution of the primary response, we found no differences in frequency of total monocytes, monocyte subsets or plasmacytoid dendritic cells among PBMCs between CoV2^+^ and HC individuals (**E.D. Fig. 2**). We also found no differences in γδ or αβ CD3^+^ T cell frequencies (CD4^+^ or CD8^+^), nor in the cell cycle status, expression of molecules associated with activation, migration, function or proportions of various CD45RA^-^ memory T cell subsets (**E.D. Fig. 3**). Together, these data demonstrate that the inflammatory response associated with acute infection had resolved by the Visit 1 time point and the early immune memory phase had commenced.

**Figure 2:**
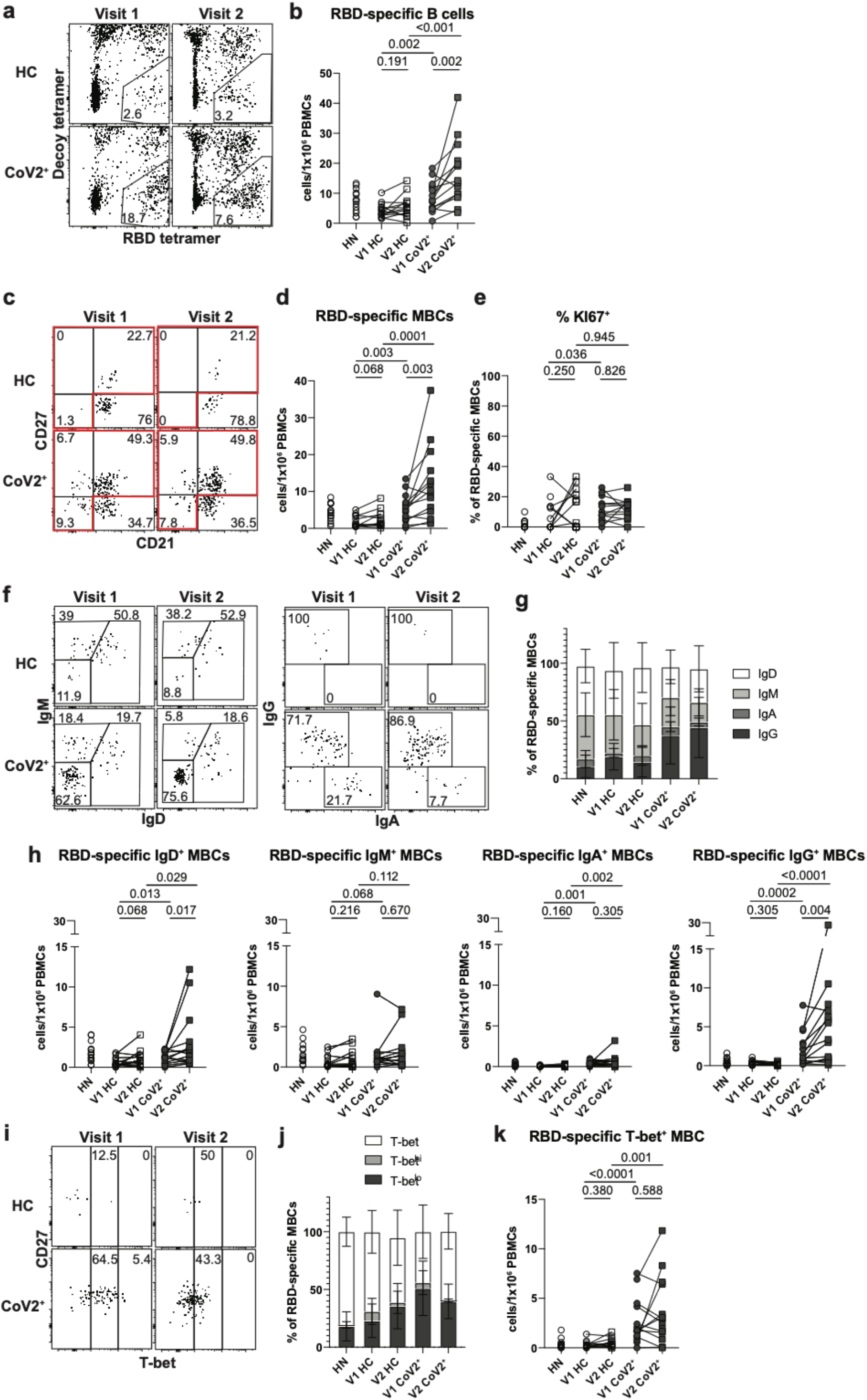
RBD-specific MBCs form and persist in PBMCs post-mild COVID-19. **a)** Representative gating of Live CD3CD14CD16^-^ cells for SARS-CoV-2 RBD-specific cells (RBD tetramer^+^Decoy^-^) and **b)** number of RBD-specific B cells (RBD tetramer^+^Decoy^-^CD20^+^) from SARS-CoV-2-recovered (CoV2^+^) and healthy control (HC) PBMCs at Visit 1 (V1) and Visit 2 (V2). Gating strategy shown in Extended Data Figure 5c. **c)** Representative gating and **d)** Number of RBD-specific memory B cells (MBCs: CD21^+^CD27^+^/CD21^-^CD27^+^/CD21^-^CD27^-^ populations outlined in red in c)(HN n=14, V1 HC n=12, V2 HC n=13, V1 CoV2^+^ n=15, V2 CoV2^+^ n=14). **e)** Frequency of cycling (Ki67^+^) RBD-specific MBCs. **f)** Representative gating, **g)** frequency (HN n=14, V1 HC n=12, V2 HC n=13, V1 CoV2^+^ n=15, V2 CoV2^+^ n=14) and **h)** number of RBD-specific MBCs expressing the BCR isotypes IgD, IgM, IgA and IgG. **i)** Representative gating, **j)** frequency and **k)** number of RBD-specific MBCs expressing T-bet. Statistical significance determined by two-tailed, Mann-Whitney test (HC vs. CoV2^+^) and twotailed Wilcoxon signed rank test (V1 vs V2). Error bars represent mean and SD (HN n=14, V1 HC n=15, V2 HC n=15, V1 CoV2^+^ n=15, V2 CoV2^+^ n=14 unless otherwise noted, 2 experiments).

**Figure 3:**
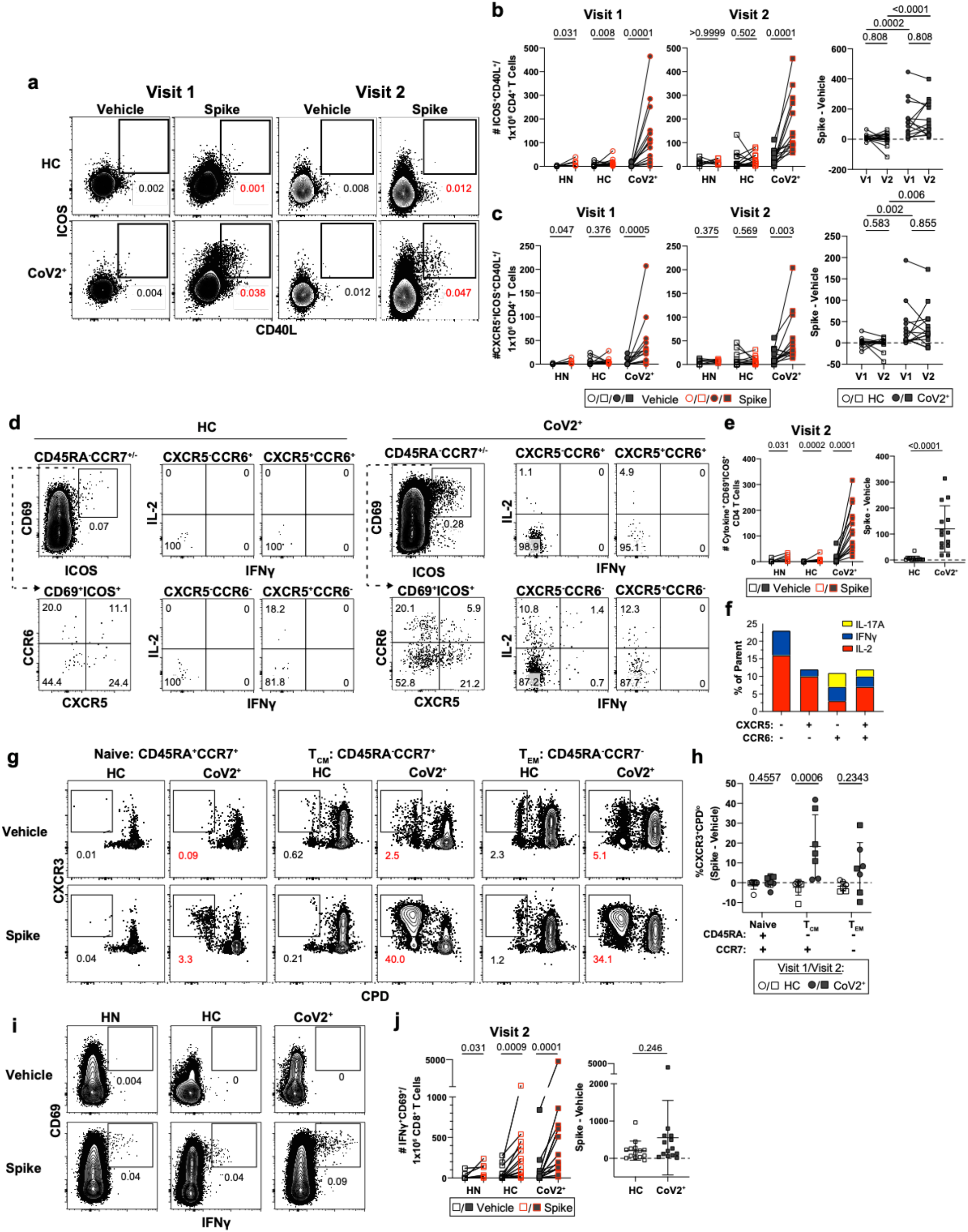
*Ex vivo* reactivation of spike-specific CD4^+^ T Cells reveals durable and functional immune memory in SARS-CoV-2-recovered individuals. **a)** Representative flow cytometry plots 20 hours after Vehicle control or Spike-stimulation of PBMCs from HC and CoV2^+^ individuals demonstrating T cell upregulation of CD40L and ICOS on CD45RA^-^CD4^+^ T cells. **b)** Enumeration of total CD40L^+^ICOS^+^ and **c)** CXCR5^+^CD40L^+^ICOS^+^ (cTfh) per 1e6 CD4^+^ T Cells and paired CoV2^+^ data from Visit 1 and Visit 2 represented as frequency of spike minus vehicle. **d)** Representative flow cytometry plots and **e)** number of CD69^+^ICOS^+^ CD4^+^ T Cells producing intracellular cytokines and number producing cytokine after incubation with spike minus number after incubation with vehicle. **f)** Relative distribution of effector cytokine production in memory T Cell compartments (CCR6^+/-^ cTfh and non-cTfh) following ex vivo stimulation for 20 hrs; (IFN-y; blue) (IL-2; red) (IL-17A; yellow) from **(d)**. **g)** Antigen-specific T cell proliferation of sorted CD4^+^ naive or memory T cells in control and CoV2^+^ PBMCs. Proliferation following 5-6 day co-culture with SARS-CoV-2 spike protein-pulsed autologous monocytes. **h)** Antigen-specific expansion represented as frequency of spike minus vehicle, CXCR3^+^CPD^low^ responding cells. **i)** Representative flow cytometry plots and **j)** quantification of spike-specific CD8^+^ T Cells in control and Cov2^+^ PBMCs stimulated with SARS-CoV-2 spike protein. **a-h)** Significance was determined by Kruskal-Wallis test correcting for multiple comparisons using FDR two-stage method. Adjusted p values are reported. **i-j)** Significance was determined by two-tailed, non-parametric Mann-Whitney tests. **a-j)** Data represented as mean and SD; Each symbol represents one donor. **a-f, i-j)** n=7 HN, n=14 HC, n=14 CoV2^+^(2 experiments). **g-h)** n=3 V1 HC, n=4 V2 HC, n=3 V1 CoV2^+^, n=4 V2 CoV2^+^ (2 experiments).

### Mild COVID-19 induces persistent, neutralizing anti-SARS-CoV-2 IgG antibody

Humoral immune responses are characterized by a first wave of short-lived, low-affinity antibody-secreting PBs followed by a subsequent germinal center (GC) response that generates high-affinity MBCs and antibody-secreting LLPCs. LLPCs can maintain detectable serum antibody titers for months to many years, depending upon the specific viral infection^13^. Thus, it is critical to distinguish the first wave of waning PB-derived antibodies from the later wave of persistent LLPC-derived antibodies that can neutralize subsequent infections, potentially for life. We therefore first determined that CD19^+^CD20^lo^CD38^hi^ PBs were no longer present at elevated frequencies in CoV2^+^ individuals relative to HCs at Visit 1 (**E.D. Fig. 4a**). Other measures of recent B cell activation in non-PB B cells include increased Ki67 expression (indicating cells have entered the cell cycle) and expression of T-bet^14^. There are small increases in both the frequencies of Ki67^+^ and T-bet^+^ B cells at the Visit 1 time point compared to HC, but not at the Visit 2 time point (**E.D. Fig. 4b,c**). These data suggest that while PBs associated with controlling acute infection are no longer detectable in CoV2^+^ individuals at Visit 1, other B cell fates are still contracting. However, by Visit 2, these B cell phenotypes have returned to homeostasis (**E.D. Fig. 4a-c**).

**Figure 4.**
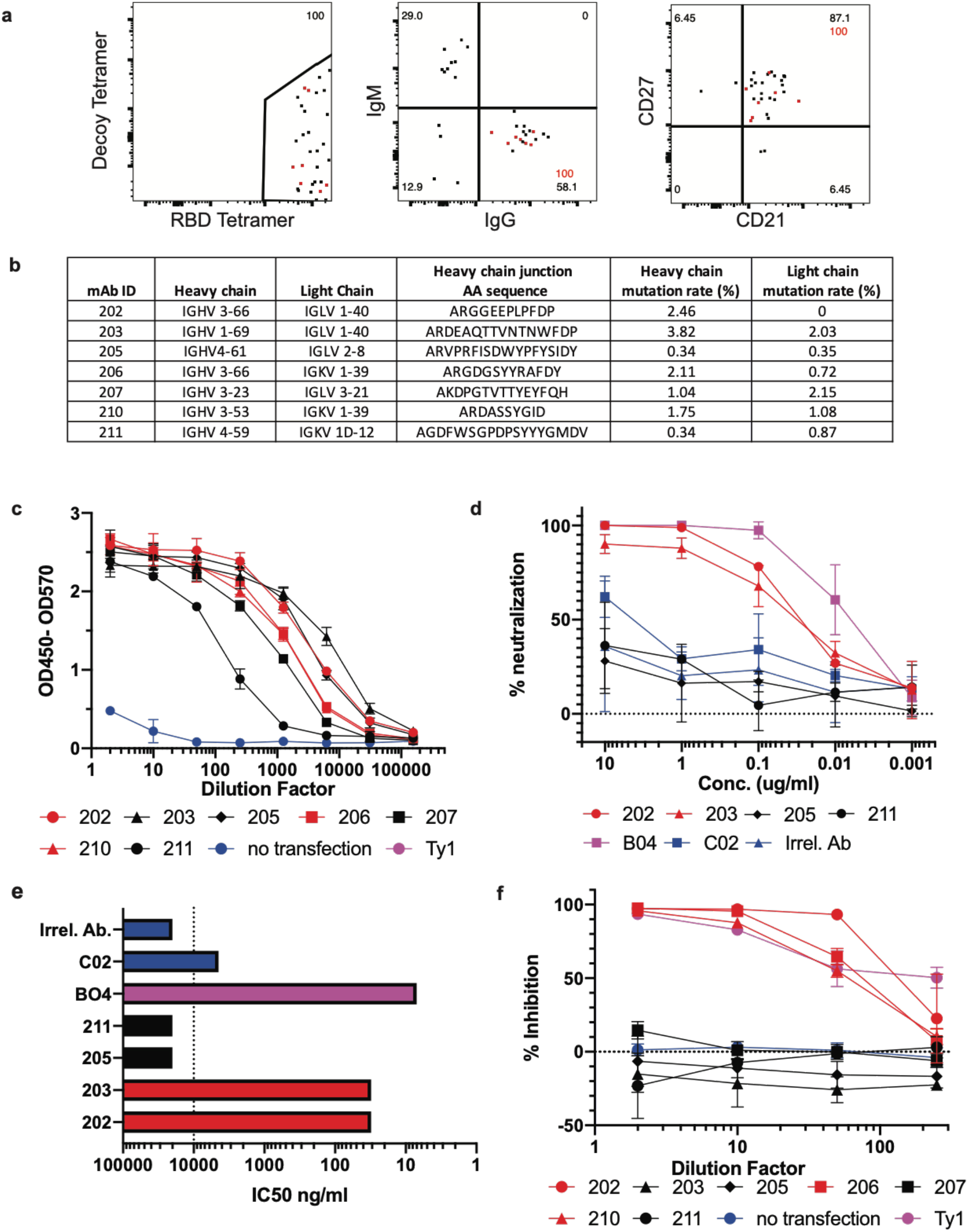
Generation of neutralizing antibodies by RBD-specific MBCs. **a**) Flow plots of index sorted RBD-tetramer specific B cells (gating scheme in Extended Data Figure 7a). B cell receptors (BCRs) cloned from cells shown in red. **b**) Heavy and light chain gene usage, somatic hypermutation rate and VDJ junction sequence of cloned BCRs. **c**) Anti-RBD ELISA of culture supernatants from cells transfected to express one of the monoclonal antibodies compared to a known RBD-binding and neutralizing antibody (Ty1) and supernatant from untransfected cells (no transfection). **d)** Neutralization capacity of purified monoclonal antibodies as measured by PRNT. BO4 and CO2 are previously identified strong and weak neutralizing murine antibodies. **e)** IC50 values of antibodies calculated from PRNT. Dotted line represents the limit of detection. **f)** Inhibition of RBD-ACE2 binding by culture supernatants from antibody transfections (antibodies with high inhibitory capacity shown in red).

Antibodies measured at Visit 1 might include contributions from short-lived plasmablasts, while those measured at Visit 2, long after PBs have contracted, represent contributions from LLPCs in the bone marrow. We therefore examined the SARS-CoV-2-specific IgG, IgM and IgA antibodies at Visit 1 and Visit 2^15^. At Visit 1, 100% of CoV2^+^ individuals had plasma anti-RBD IgG levels 3 SDs above the mean of HCs, as measured by ELISA area under the curve (AUC), in accordance with studies showing 100% seroprevalence by day 14^10^ (**Fig. 1b**). Additionally, 93% of CoV2^+^ individuals had anti-RBD IgM and 73% had anti-RBD IgA above this negative threshold. Almost all CoV2^+^ individuals possessed IgG (100%), IgM (100%), and IgA (93%) anti-spike antibodies above the threshold at Visit 1 as well (**E.D. Fig. 4d**). Levels of anti-RBD and anti-spike binding were highly correlated for all isotypes (**E.D. Fig. 4e**). At Visit 2, all CoV2^+^ individuals maintained anti-RBD IgG levels above the negative threshold and 71% and 36% had maintained anti-RBD IgM and IgA, respectively (**Fig. 1b**). Anti-RBD IgG levels decreased only slightly among CoV2^+^ individuals between time points and 36% of CoV2^+^ individuals had the same or increased levels at Visit 2. Anti-RBD IgM and IgA, however, decreased substantially from Visit 1 to Visit 2 (**Fig. 1c, E.D. Fig. 4f**).

As spike protein, and specifically the RBD, is key for viral entry into the cell, antibodies that target the RBD can be potent inhibitors of infection^8,9^. To determine whether CoV2^+^ individuals form and maintain neutralizing antibodies, we tested for SARS-CoV-2 neutralization indirectly using a cell-free competition assay (surrogate virus neutralization test, sVNT) and directly in a plaque reduction neutralization test (PRNT)^16^. CoV2^+^ plasma inhibited RBD binding to ACE2 significantly more than HC plasma by sVNT and RBD inhibition correlated strongly with anti-

RBD IgG levels at both time points (**Fig. 1d,e**). Further, RBD inhibition capacity was maintained or increased in the majority of CoV2^+^ individuals from Visit 1 to Visit 2 (**Fig. 1f, E.D. Fig. 4g**). Neutralization by PRNT correlated strongly with RBD inhibition at both time points (**Fig. 1g, E.D. 4h**) and was similarly maintained between visits (**Fig. 1h**). By the latest time point in our study, 86% of CoV2^+^ individuals still had better RBD-inhibiting plasma than HCs and 71% had better neutralizing plasma (measured as above HC mean + 3 SDs). These data are consistent with the emergence of predominantly IgG^+^ RBD and spike-specific LLPCs that maintain detectable neutralizing anti-SARS-CoV-2 antibody to at least 3 months post-symptom onset.

### Mild COVID-19 induces a sustained enrichment of RBD-specific memory B cells

The presence of SARS-CoV-2-neutralizing antibodies three months post-symptom onset in CoV2^+^ individuals suggests GC-derived memory LLPCs have formed. GC-derived MBCs play a critical role in the formation of antibody secreting cells upon antigen re-exposure. Therefore, we tested whether SARS-CoV-2-specific MBCs were also formed and maintained in CoV2^+^ individuals throughout the study time course. We generated RBD tetramer reagents and used enrichment strategies to identify rare RBD-specific cells that are otherwise undetectable in bulk assessments^17^. We confirmed specificity in RBD immunized mice and then used the RBD-tetramer to identify, enumerate and phenotype rare, RBD-specific B cells in our HN, HC and CoV2^+^ individuals (**E.D. Fig. 5a,b**; **Fig. 2a**). Gates used to phenotype RBD-specific B cells were defined on total B cell populations (**E.D. Fig. 5c**). At Visit 1, RBD-specific B cells were significantly expanded in CoV2^+^ individuals compared to HCs and their numbers were increased further at Visit 2 **(Fig. 2a, b)**. The proportion and number of RBD-specific MBCs (defined by CD21 and CD27 expression) in CoV2^+^ samples was significantly greater than in HCs and increased from Visit 1 to Visit 2 (**Fig. 2c,d, E.D. Fig. 5d**). While RBD-specific B cells in HN samples had a similar proportion of MBCs as in CoV2^+^ samples, they contained substantially fewer cells. In addition, RBD-specific MBCs were largely quiescent with very few expressing Ki67 **(Fig. 2e, E.D. Fig. 5e)**. MBCs expression of class-switched B cell receptors (BCRs) is another marker of GC-derivation. We therefore assayed BCR isotype expression on RBD-specific MBCs and found enriched populations of IgA- and IgG-expressing MBCs in CoV2^+^ individuals at both time points **(Fig. 2f-h, E.D. Fig. 5f)**. Of note, while small numbers of RBD-specific MBCs were detected in controls, these cells were predominantly unswitched (IgM^+^ and IgD^+^), suggesting they may represent cross-reactive MBCs possibly generated in response to one of the human coronaviruses that cause 15% of common colds^18–20^.

An additional measure of antiviral MBC function is the graded expression of T-bet^14^. MBCs that express low-levels of T-bet are associated with rapid differentiation into secondary PBs that produce high affinity, viral-specific antibodies during a secondary infection^21^. We found a higher proportion and number of T-bet^+^, and specifically T-bet^lo^, RBD-specific MBCs in CoV2^+^ individuals compared with HCs at Visit 1 and the higher numbers were maintained at Visit 2 (**Fig. 2i-k, E.D. Fig. 5g,h**). T-bet^hi^ MBCs are considered to be recently activated and often found enriched during chronic infection^21^. Consistent with SARS-CoV-2 being an acute infection^11^, we found very few RBD-specific T-bet^hi^ MBCs in CoV2^+^ individuals at either memory time point (**E.D. Fig. 5i**). Our data demonstrate that SARS-CoV-2 infection induces the generation of RBD-specific Tbet^lo^IgG^+^CD21^+^CD27^+^ “classical” MBCs likely derived from a GC^22^. Furthermore, numbers of these MBCs were not only maintained, but increased from one to three months post-symptom onset.

### SARS-CoV-2 infection induces durable, functional spike-reactive CD4^+^ T cells

The presence of T-bet^+^ RBD-specific MBCs suggested that antigen-specific memory T cell responses were also likely to be elicited in CoV2^+^ individuals. To enumerate SARS-CoV-2-specific memory T cells, total PBMCs from control or CoV2^+^ individuals were incubated with spike protein and expression of activation markers was assessed (**Fig. 3a**)^23,24^. PBMCs from CoV2^+^ individuals at Visit 1 and 2 displayed robust re-activation of spike-specific CD4^+^ memory T responses, as measured by increased expression of ICOS and CD40L (two molecules associated with B cell help upon re-activation), while PBMCs from HC and HN individuals did not (**Fig. 3a,b**). There were no significant differences in the numbers of responding cells in CoV2^+^ individuals between the two visits, suggesting spike-specific memory CD4^+^ T cells were maintained throughout the study (**Fig. 3b**). Furthermore, greater numbers of CXCR5-expressing circulating T follicular helper (cTfh) cells^25^, which provide B cell help, were found within the population of S-specific ICOS^+^CD40L+CD4^+^ cells in CoV2^+^ individuals than in healthy controls at both visits (**Fig. 3c**). Together these data suggest that SARS-CoV-2-specific memory CD4^+^ T cells maintain the capacity to provide B cell help even at three months post-symptom onset.

Memory CD4^+^ T cells produce cytokines within hours of activation, whereas naive T cells take days^26^. We first examined cytokine production from activated CD4^+^ memory CXCR5-non-Tfh cells and CXCR5^+^ cTfh cells identified in the assay above (**Fig. 3b)**. S-specific CCR6^+^CXCR5^+^ cTfh cells, associated with IL-17 production, and a smaller population of CXCR3^+^CXCR5^+^ cTfh cells, associated with IFNγ production, were recently described in a predominantly mild to moderate cohort 30 days post symptom onset ^27^ We therefore analyzed activated ICOS^+^CD69^+^ S-specific cells for expression of CCR6 and CXCR5 and then cytokine expression was examined in each population based on gating on a PMA positive control (**Fig 3d, E.F. 6a)**. Although multiple cytokines associated with Tfh function were assessed, only IFNγ, IL-17 and IL-2 cytokine producing cells were significantly expressed in activated S-specific memory CD4^+^ cells in CoV2^+^ individuals compared to HCs (**Fig. 3d-f**). Small numbers of S-specific cells were measured in HCs after stimulation compared to vehicle alone that reflect previously described S-specific cross-reactivity^20,28^, but far greater responses were seen in the CoV2^+^ individuals (**Fig. 3e)**. Three months post symptom onset we found a higher frequency of CCR6-cTfh cells that produced Th1 cytokines, IFNγ and IL-2, suggesting a dominant Th1 response in CoV2^+^ individuals (**Fig. 3f**).

To further define the types of antigen-specific CD4^+^ memory T cells in CoV2^+^ individuals without relying on secretion of specific cytokines, we assessed memory CD4^+^ T cell proliferation in response to spike restimulation. For this, we sorted CD45RA^+^ naive, CD45RA^-^CCR7^+^ central memory (Tcm) and CD45RA^-^CCR7^+^ effector memory (Tem) T cells from HC or CoV2^+^ individuals (**E.D. Fig. 6b**), then measured the proliferative capacity of each sorted population following culture with autologous CD14^+^ monocytes and recombinant spike protein (**Fig. 3g, h; E.D. Fig 6c**). Only Tcm cells from CoV2^+^ individuals taken at both Visit 1 and Visit 2 displayed significant proliferation frequencies compared to HC samples, although substantial proliferative responses by Tem cells were observed in some CoV2^+^ individuals (**Fig. 3h**). We also examined the expression of CXCR3 and CCR6 on S-specific, proliferated memory cells and found that the majority of cells that had proliferated, as measured by the dilution of cell proliferation dye (CPD^lo^) expressed CXCR3, in keeping with Type 1 cytokine production in the previous assay. Spike-specific Tcm, and potentially Tem, are therefore maintained throughout our study and have the ability to proliferate and re-populate the memory pool upon antigen re-encounter.

While much recent work has focused on antibodies and B cells, memory CD8^+^ T cells are uniquely positioned to kill virus infected cells through their directed expression of cytokines and cytolytic molecules. S-specific memory CD8^+^ T cells that persisted for three months after mild COVID-19 disease could be identified by expression of the activation marker CD69 and the cytokine IFNγ after overnight stimulation with spike **(Fig 3i)**. Unlike CD4^+^ memory T cells, activated cytokine-expressing CD8^+^ T cells were significantly increased over vehicle controls in both control and CoV2^+^ groups **(Fig. 3j)**. Together, these data demonstrate that both CD4^+^ and CD8^+^ SARS-CoV-2-specific memory T cells are maintained and are able to produce effector cytokines after restimulation three months post-symptom onset in mildly symptomatic COVID-19 individuals.

### Mild COVID-19-induced SARS-CoV-2-specific MBCs can express neutralizing antibodies

Since SARS-CoV-2 RBD-specific MBC and S-specific CD4^+^ cTfh were enriched in CoV2^+^ individuals after 3 months, we assessed whether these MBCs could produce neutralizing antibodies if they were reactivated by a secondary infection. To this end, we index sorted single RBD-specific B cells and sequenced the BCRs from 3 CoV2^+^ individuals at Visit 1 (**E.D. Fig. 7a**). Of the class-switched (IgG^+^) RBD-specific classical MBCs (CD21^+^CD27^+^) we sorted, we randomly selected 7 to be cloned and expressed as IgG1 monoclonal antibodies (**Fig 4a**). This set of antibodies utilized a wide variety of heavy and light chains, had all undergone somatic hypermutation and were all unique clones (**Fig. 4b**). These antibodies were first expressed in small scale cultures. Transfection supernatants were assessed for antibody expression by IgG ELISA (**E.D. Fig 7b**) and specificity by RBD ELISA where all 7 showed strong binding to RBD (**Fig. 4c**). The first 4 antibodies cloned were expressed on a larger scale and purified. The specificity of these purified antibodies for RBD was again confirmed by ELISA (**E.D. Fig. 7c**) and their ability to prevent SARS-CoV-2 infection was tested via PRNT assay. Two of the four tested (#202 and 203) showed strong virus neutralization (**Fig. 4d**), with IC50 values of 31 ng/ml for both (**Fig. 4e**). This was comparable to a previously published strongly neutralizing mouse antibody (B04) which was included as a positive control (IC50=7ng/ml)^29^. Two of the RBD-specific antibodies were unable to inhibit virus infection, similar to a non-neutralizing mouse antibody (C02) and an irrelevant *Plasmodium*-*specific* human antibody. Three more monoclonal antibodies in addition to the 4 above were assessed for their capacity to inhibit RBD binding to the ACE2 receptor by sVNT assay (**Fig. 4f**). Three of the seven were able to inhibit RBD binding to ACE2, similarly to a strongly neutralizing alpaca nanobody^30^. Interestingly, #203, which neutralized live virus, did not inhibit binding in this assay, while #202 both inhibited binding and neutralized the virus. Overall 50% of the antibodies tested showed inhibitory activity by one or both of these methods. Thus, RBD-specific MBCs induced by SARS-CoV-2 infection are capable of producing neutralizing antibodies against the virus and could thus contribute to protection from a second exposure to SARS-CoV-2.

### Discussion

In the absence of a vaccine, natural infection-induced herd immunity could play a key role in reducing infections and deaths. For this to be possible, individuals that experience mild COVID-19 would need to develop and sustain protective immune memory. Here, we found that individuals that recovered from mildly symptomatic COVID-19 had an expanded arsenal of SARS-CoV-2-specific immune mediators: neutralizing antibodies, IgG^+^T-bet^lo^ classical MBCs, circulating cytokine-producing CXCR5^+^ Tfh1 cells, proliferating CXCR3^+^ CD4^+^ memory cells and IFNγ producing CD8^+^ T cells that were maintained to at least three months post-symptom onset. This study predicts that these recovered individuals will be protected from a second SARS-CoV-2 infection and, if so, suggests that Th1 memory should be the target of vaccine elicited memory.

Although long-lived immune memory can form to most viruses, some studies examining the longevity of the response to coronaviruses have suggested that this is not the case^31–33^. However, more recent studies, including our own, have examined memory time points when only LLPCs, and not short-lived PBs, are producing circulating antibodies. Our study, along with three others clearly demonstrates elevated IgG^+^ RBD-specific plasma antibodies and neutralizing plasma are generated and maintained for at least 3 months post-SARS-CoV-2 infection^34–36^.

While antibodies reveal the contributions of LLPCs, functional virus-specific memory B and T cells can also be key to protective immune memory^37^. Although previous studies have measured the emergence of SARS-CoV-2-specific MBCs within a month of infection ^27,38^, we characterized SARS-CoV-2-specific MBCs at one and three months from symptom onset. Our study revealed a prominent population of RBD-specific IgG^+^CD27^+^CD21^+^T-bet^lo^ MBCs, which has been associated in other infections with rapid differentiation into antibody-secreting PBs upon re-exposure^5^, effective antiviral responses^39^and long-lived protection^3^. Furthermore, we found some of the RBD-specific MBCs at Visit 1 expressed BCRs capable of neutralizing the virus when expressed as antibodies. Since the numbers of these IgG^+^ RBD-specific MBCs were not only sustained, but continued to increase between one and three months, we predict they are GC-derived. Thus, MBCs at three months would have undergone increased affinity maturation and we would expect an even higher percentage will be capable of producing neutralizing RBD-specific antibodies upon re-infection.

MBC reactivation requires interactions with memory CD4^+^ T cells, which reactivate MBCs through their expression of key molecules associated with T-B interactions including CXCR5, ICOS, CD40 and a variety of cytokines. SARS-CoV-2-specific CD4^+^ memory T cells in recovered individuals exhibited the capacity to express all of these molecules and to undergo robust proliferation upon re-exposure to spike protein. Notably, S-specific CD4^+^ memory T cells from CoV2^+^ individuals rapidly displayed increased levels of ICOS and CD40L on CXCR5^+^ and CXCR5^-^ cells after stimulation as well as expression of Th1^-^ and Th17^-^ associated cytokines. These results are consistent with another recent report of SARS-CoV-2- specific cTfh cells^27^, although they detected a high frequency of Th17-like cTfh cells, which could be due to the earlier time point they were examining as Th17 cells can develop into Th1 cells late in an immune response^40^. The expression of IFNγ and IL-17 by these cells is notable as these cytokines are associated with class-switching to IgG and IgA isotypes, respectively ^41,42^. We also found crossreactive memory B and T cells in healthy controls, as has been previously noted^43^. It is difficult to measure their contribution to the expanded populations of SARS-CoV-2-specific cells we found in our CoV2^+^ cohorts, and therefore impossible to evaluate their protective capacity. However, we can conclude that mild COVID-19 induces an expanded population of functionally diverse memory lymphocytes compared to the cross-reactive pool present in our controls.

Studies of reinfection have yet to be done in humans, but macaques infected with SARS-CoV-2 were protected from rechallenge^44^. This further suggests that the immune memory induced by mild COVID-19 that we observed will be protective. While additional studies are needed to understand variability of responses in a larger cohort and to determine how long memory to SARS-CoV-2 infection is truly maintained, our work suggests that mild COVID-19 induces persistent immune memory poised for a coordinated, protective response to re-exposure that could contribute to herd immunity and curtailing this pandemic.

## Data Availability

All data generated or analysed during this study are included in this published article (and its supplementary information files) or available from the corresponding author upon reasonable request.

## Methods

### Ethics Statement

This study was approved by the University of Washington Institutional Review Board (Gale Lab, IRB 00009810). Informed consent was obtained from all enrolled participants. Samples were deidentified prior to transfer to the Pepper Lab.

### Study Participants

The study was conceptualized utilizing a prospective case-control design. Cases and controls were identified from a cross-sectional cohort study that recruited via print and online advertising from the Seattle metropolitan area (**E.D. Table 1**). Cases (n=15) were selected based on a reported history of a positive SARS-CoV-2 PCR nasal swab. Controls (n=17) were selected based on having no prior positive SARS-CoV-2 PCR nasal swab and having no detectable SARS-CoV-2 RBD- or S-specific IgG or IgM plasma antibodies (within mean + 3 SD of 5 de-identified plasma samples drawn prior to 2020 generously donated by Wesley C. Van Voorhis). At the time of enrollment, information was collected from all participants regarding recent illness symptoms and severity. All CoV2^+^ cases reported at least one symptom but all were classified as mild disease, as none required hospitalization. Historical negative control PBMCs (n=14) were sourced from the BRI PBMC repository. Samples were drawn prior to 2020 and age and sex matched to the CoV2^+^ cases.

### Peripheral blood mononuclear cell (PBMC) and plasma collection

6-10 milliliters of venous blood from study volunteers were collected in EDTA tubes and spun at 1500xg for 10 minutes. Plasma was collected, heat-inactivated at 56°C for 30 minutes, aliquoted and stored at −80°C. The cellular fraction was resuspended in PBS and PBMC were separated from RBC using Sepmate PBMC Isolation Tubes (STEMCELL Technologies) according to manufacturer’s instruction and frozen at −80°C before being stored in liquid nitrogen. PBMCs were thawed at 37C and washed twice before use.

### SARS-CoV-2 Protein Production and Purification

#### Plasmid construction

The SARS-CoV-2 S^B^ (BEI NR-52422) construct was synthesized by GenScript into pcDNA3.1- with an N-terminal mu-phosphatase signal peptide and a C-terminal octa-histidine tag (GHHHHHHHH). The boundaries of the construct are N-_328_RFPN_331_ and C-_528_KKST_531_. The SARS-CoV-2 S-2P ectodomain trimer (GenBank: YP_009724390.1, BEI NR-52420; cite PMID 32155444) was synthesized by GenScript into pCMV with an N-terminal mu-phosphatase signal peptide and a C-terminal TEV cleavage site (GSGRENLYPQG), T4 fibritin foldon (GGGSGYIPEAPRDGQAYVRKDGEWVLLSTPL), and octa-histidine tag (GHHHHHHHH). The construct contains the 2P mutations (proline substitutions at residues 986 and 987; PMID 28807998) and an 682SGAG685 substitution at the furin cleavage site. A pCAGGS vector containing the spike protein RBD from SARS-CoV-2 (Wuhan-Hu-1 isolate) was generously provided by Florian Krammer.

#### Transient expression

Constructs were produced in Expi293F cells grown in suspension using Expi293F expression medium (Life technologies) at 33°C, 70% humidity, and 8% CO2 rotating at 150 rpm. The cultures were transfected using PEI-MAX (Polyscience) with cells grown to a density of 3.0 million cells per mL and cultivated for 3 days. Supernatants was clarified by centrifugation (5 minutes at 4000 rcf), addition of PDADMAC solution to a final concentration of 0.0375% (Sigma Aldrich, #409014), and a second spin (5 minutes at 4000 rcf).

#### Purification of His-tagged proteins

Proteins were purified from clarified supernatants via a batch bind method where each supernatant was supplemented with 1 M Tris-HCl pH 8.0 to a final concentration of 45 mM and 5 M NaCl to a final concentration of ~310 mM). Talon cobalt affinity resin (Takara) was added to the treated supernatants and allowed to incubate for 15 minutes with gentle shaking. Resin was collected using vacuum filtration using a 0.2 μm filter and transferred to a gravity column. The resin was washed with 20 mM Tris pH 8.0, 300 mM NaCl, and the protein was eluted with three column volumes of 20 mM Tris pH 8.0, 300 mM imidazole, 300 mM NaCl. The batch bind process was then repeated and the first and second elutions combined. SDS-PAGE was used to assess purity. Purified S-2P trimer was concentrated to ~1 mg/mL and dialyzed into 50 mM Tris pH 8, 150 mM NaCl, 0.25% L-Histidine, 5% glycerol in a hydrated 10k molecular weight cutoff dialysis cassette (Thermo Scientific). The purified RBD protein was dialyzed into 50 mM Tris pH 7, 185 mM NaCl, 100 mM Arginine, 4.5% glycerol, 0.75% w/v CHAPS. Due to inherent instability, S-2P was immediately flash frozen and stored at −80°C.

### Tetramer generation

Recombinant trimeric spike and the RBD domain were both biotinylated using a EZ-Link Sulfo-NHS-LC Biotinylation Kit (ThermoFisher), tetramerized with streptavidin-PE (Agilent) and stored in 50% glycerol at −20°C as previously described^17^. Decoy reagent was generated by tetramerizing an irrelevant biotinylated protein with SA-PE previously conjugated to AF647 using an Alexa Fluor 647 Antibody Labeling Kit (ThermoFisher).

### Tetramer validation in mice

Adult C57BL/6j mice (The Jackson Laboratory) were immunized with 50ug SARS-CoV-2 RBD in CFA in the footpad and, 7-10 days later, popliteal lymph nodes were dissected, mashed and stained in 10nM decoy-PE-APC tetramer and then 10nM RBD-PE as described^17^and as described below for immunophenotyping B cells. Cells stained for surface markers as indicated **(Supplemental Table 1)** and run on the LSRII (BD). Data analyzed with FlowJo10 (Becton Dickinson). Mice were purchased from The Jackson Laboratory and maintained under specific pathogen free conditions at the University of Washington. All mouse experiments were performed in accordance with the University of Washington Institutional Care and Use Committee guidelines.

### ELISA

96-well plates (Corning) were coated with 2 ug/mL of recombinant SARS-CoV-2 RBD or trimeric spike protein diluted in PBS and incubated at 4°C overnight. Plates were washed with PBS-T (PBS containing 0.05% Tween-20) and incubated with blocking buffer (PBS-T and 3% milk) for 1 hour at room temperature (RT). Serum, culture supernatants or monoclonal antibodies were serially diluted in dilution buffer (PBS-T and 1% milk) in triplicate, added to plates, and incubated at RT for 2 hours. Secondary antibodies were diluted in dilution buffer as follows: anti-human IgG-HRP (Jackson ImmunoResearch) at 1:3000, anti-human IgM-HRP (Southern Biotech) at 1:3000, or anti-human IgA-HRP (Southern Biotech) at 1:1500. Plates were incubated with secondary antibodies for 1 hour at RT, then detected with 1X TMB (Invitrogen) and quenched with 1M HCl. Sample optical density (OD) was measured by a spectrophotometer at 450nm and 570nm. CR3022, a human SARS-CoV antibody previously determined to cross-react with SARS-CoV-2 was used as a positive control. IgG in culture supernatants was measured using a Human IgG ELISA Kit (Stemcell) according to the manufacturer’s instructions. Data was analysed in Prism (GraphPad).

### Receptor-binding inhibition assay

96-well plates (Corning) were coated with 5 ug/mL of recombinant human ACE2-Fc diluted in 100mM carbonate-bicarbonate buffer (pH 9.6) and incubated at 4°C overnight. Plates were washed with PBS-T and incubated with blocking buffer for 1 hour at RT. Plasma or monoclonal antibody supernatants were serially diluted in triplicate in dilution buffer and incubated with 18ng of recombinant SARS-CoV-2 RBD-HRP (conjugated using Abcam HRP conjugation kit) for 1 hour at 37C. Blocked plates were washed and incubated with the pre-incubated serum and RBD-HRP for 1 hour at RT, then detected with TMB and 1M HCl. OD was measured by a spectrophotometer at 450nm and 570nm. RBD-HRP alone and serum with no RBD-HRP incubation were used as controls. The % inhibition was calculated as (1 - Sample OD value/Average Negative Control OD value) x 100. Data was analysed in Prism (GraphPad).

### Plaque reduction neutralization test (PRNT)

PRNT assays were performed as previously described^45^. Briefly, heat inactivated plasma was diluted 1:5 followed by four 4-fold serial dilutions and monoclonal antibodies were diluted 1:10 followed by 4 10-fold serial dilution and mixed 1:1 with 600 PFU/ml SARS-CoV-2 WA-1 (BEI resources) in PBS+0.3% cold water fish skin gelatin (Sigma). After 30 minutes of incubation at 37°C, the plasma/virus mixtures were added to 12 well plates of Vero cells and incubated for 1 hour at 37C, rocking every 15 minutes. All dilutions were done in duplicate, along with virus only and no virus controls. Plates were then washed with PBS and overlaid with a 1:1 mixture of 2.4% Avicel RC-591 (FMC) and 2X MEM (ThermoFisher) supplemented with 4% heat-inactivated FBS and Penicillin/Streptomycin (Fisher Scientific.) After a 48 hour incubation, the overlay was removed, plates were washed with PBS, fixed with 10% formaldehyde (Sigma-Aldrich) in PBS for 30 minutes at room temp and stained with 1% crystal violet (Sigma-Aldrich) in 20% EtOH. % Neutralization was calculated as (1 - # sample plaques/# positive control plaques) x 100. Data was analysed in Prism (GraphPad) and IC50 was calculated by sigmoidal interpolation method.

### Cell Enrichment, Stimulations and Flow Cytometry

#### Immunophenotyping and sorting RBD-specific B cells

Thawed PBMCs were first stained with Decoy tetramer and then with RBD tetramer prior to incubation with anti-PE magnetic beads and magnetic bead enrichment (Miltenyi Biotec) as previously described.^17^ Bound cells were stained with surface antibodies **(SI Table 1)** and, if required, were fixed/permeabilized using eBioscience FoxP3 Fix/Perm kit (ThermoFisher; 005521-00) for 30 minutes, followed by incubation with intracellular antibodies **(SI Table 1)**. Stained samples were run on a LSRII flow cytometer and analyzed using FlowJo (Becton Dickinson). For B cells sorting experiments, single tetramer-specific B cells were indexed sorted on a FACSAriaII cell sorter and collected in a 96-well PCR plate containing SMART-Seq v4 capture buffer (Takara Bio).

#### Immunophenotyping of PBMCs

For surface phenotyping, total PBMCs (innate cells) or PBMCs from the negative fraction of the antigen-specific B Cell magnetic columns (for lymphocytes) were washed and incubated with fluorescently conjugated antibodies. Staining for cTfh analyses were performed as follows: chemokine-receptors and transcription factors (40 minutes, RT), surface antigens (20 minutes, 4°C)**(SI Table 1)**. Intracellular staining was performed using eBioscience FoxP3 Fix/Perm kit (ThermoFisher; 00-5521-00)**(SI Table 1)**. For detection of intracellular cytokine production, PBMC were stimulated with 50 ng/ml phorbol 12-myristate 13-acetate (Sigma-Aldrich) and 1 μg/ml Ionomycin (Sigma-Aldrich; I06434) with 10 μg/ml Brefeldin A (Sigma-Aldrich; B6542) and 1x dose GolgiStop/monensin (Becton Dickinson; 554724) for 4 hours. Permeabilization and fixation was performed using Cytofix/Cytoperm (Becton Dickinson; RUO 554714). Intracellular stains were performed for 30 minutes at 4°C **(SI Table 1)**. Flow cytometry analysis of innate immune populations was done on 0.5-1 million PBMCs before fraction isolation. Data was acquired on a Cytek Aurora or BD LSR Fortessa and analyzed using FlowJo10 software (Becton Dickinson).

#### Ex-Vivo spike Protein Stimulation of Peripheral Blood T Cells

PBMCs from the negative fraction of antigen-specific B Cell magnetic columns were washed and resuspended to 4×10^6^ cells/mL with complete RPMI with 10mM HEPES (ThermoFisher; 22400097) supplemented with 10% FBS, 2Me, Pen-Strep, and L-Glutamine. Spike-stimulated PBMCs were incubated with 2ug/mL full-length recombinant spike protein resuspended in PBS + 5% glycerol. Unstimulated controls received equivalent volume of PBS + 5% glycerol vehicle. Both conditions were left for 20 hours at 37C, 5-8% CO2, with addition of 10 μg/ml Brefeldin A (Sigma-Aldrich; B6542) and 1x dose GolgiStop/monensin (Becton Dickinson; 554724) for the final 5 hours to allow for intracellular detection of cytokines. Positive controls were stimulated with PMA/Ionomycin (see above) for 5 hours in the presence of Brefeldin-A and Monensin. Staining was performed as follows: chemokine-receptors (40 minutes, RT), surface antigens and cytokines (20 minutes, 4°C) **(SI Table 1)**. Cells were run on the Cytek Aurora and analyzed using FlowJo (Becton Dickenson).

#### Antigen-specific T cell proliferation

Starting with PBMC from healthy control or CoV2^+^ individuals, cell proliferation dye (CPD)- labeled, 1.25uM (ThermoFisher; 65-0840-85), sorted naive or memory T cell subsets (5 x 10^4^) were co-cultured in round-bottomed 96-well plates with irradiated autologous monocytes (5000 rads, 5 x 10^4^), and provided either full-length recombinant human spike protein (2.5ug/mL) resuspended in 5% PBS-glycerol or vehicle control. Cultures were supplemented with 5U/mL recombinant human IL-2 (Biolegend; 589104). Cellular proliferation was assessed after 5-6 days by flow cytometry **(SI Table 1)** as above and analyzed using FlowJo10 (Becton Dickenson). The percentage of CXCR3^+^CPD^lo^ cells (defined as cells that had undergone 3 or more divisions) represented as Spike - Vehicle is calculated by subtracting the vehicle control proliferation from spike-treated proliferation.

### Monoclonal antibody generation

#### BCR sequencing and cloning

Amplification of cDNA was performed using SMART-Seq v4 (Takara Bio) at half reaction volume for each sorted cell. BCR chains were amplified in a multiplex PCR using half reactions of DreamTaq (Thermo Fisher) and 1.25 ul of resulting cDNA with 3’ primers for constant regions of IgM, IgA, (5’-GGAAGGAAGTCCTGTGCGAGGC-3’, 5’-GGAAGAAGCCCTGGACCAGGC-3’, Wardemann and Busse, 2019) IgG, IgK, IgL (5’- TCTTGTCCACCTTGGTGTTGCT’-3’, 5’-GTTTCTCGTAGTCTGCTTTGCTCA-3’, 5’- CACCAGTGTGGCCTTGTTGGCTTG-3’, Smith et al, 2009) and a 5’ primer for the template switch sequence (5’-GTGGTATCAACGCAGAGTACATGGG-3’). Thermocycler conditions were 95 °C for 2 min, 30 cycles of 95 °C for 30s, 57 °C for 30s and 72 °C for 1 min.. Resulting PCR products were cleaned using 5 ul of PCR reaction, 1 ul FastAP (Thermo Fisher), and 0.5 ul Exonuclease I (ThermoFisher) for 30 minutes at 37°C and inactivated at 75°C for 15 minutes. Sanger sequencing for each purified sample was performed using each 3’ primer from the previous BCR PCR amplification. Sequences were trimmed at Q30 using Geneious and submitted to IMGT/HighV-QUEST for alignment (Alamyar et al, 2012). Primers were designed using 5’ and 3’ cDNA sequence for In-Fusion Cloning Kit and performed according to manufacturer’s instructions. If a 5’ or 3’ sequence was missing, then the closest matching IMGT germline sequence was used for primer design. Heavy chains were inserted into IgG1 vectors, kappa and lambda chains were cloned into vectors with their respective constant regions(Smith et al. 2009). Cloned plasmids were sequenced and screened by ensuring sequences of chains matched original cDNA sequence.

#### Expression and purification

For small scale transfections, 12 well plates of 293T cells at 80% confluence were transiently transfected with 0.5ug each of heavy and light chain vectors using polyethylenimine (PEI). After 16 hours, media was removed and replaced with serum-free media. After 3-4 days, supernatants were harvested and cell debris was removed by centrifugation at max speed in a microcentrifuge for 1 minute. For large scale transfections, expression vectors containing paired heavy and light chains were transiently transfected into 293T cells using polyethylenimine (PEI). Expression of recombinant full-length human IgG monoclonal antibodies were carried out in serum-free basal medium (Nutridoma-SP, Sigma-Aldrich). Four days after transfection, cell culture medium was collected and protein was purified using HiTrap™ Protein G HP column (1ml, GE Healthcare). Final IgG proteins were concentrated and buffer exchanged into 1x PBS using Millipore concentrator (30K MWCO). IgG protein concentration is determined by Nanodrop 2000 spectrophotometer.

### Statistics

Statistics used are described in figure legends and were determined using Prism (Graphpad). All measurements within a group in a panel are from distinct samples except technical replicates used in ELISAs as described. Statistical significance of all pairwise comparisons was assessed by twotailed nonparametric tests; Mann-Whitney for unpaired data and Wilcoxon signed rank tests for paired data unless otherwise noted. No multiple hypothesis testing was applied and exact p-values are displayed.

### Data Availability

All data generated or analysed during this study are included in this published article (and its supplementary information files) or available from the corresponding author upon reasonable request with the exception of a few blood draw samples that were used up in this study.

## Acknowledgements

We thank David M. Koelle for PBMC samples used for reagent testing, Florian Krammer for vectors used to express the SARS-CoV-2 S RBD, Wesley C. Van Voorhis for historical negative serum samples, Ben Murrell and the CoroNAb consortium for providing the Ty1 antibody, Ali Ellebedy for providing the BO4 and CO2 antibodies, Mike Murphy and Deleah Pettie for assistance with protein production, the Benaroya Research Institute biorepository for historical negative PBMC samples, Noah Simon for assistance with statistical analyses, Makala Hale for help with BCR sequencing, Brian Hondowicz for technical help, the Pepper, Gale, Campbell, Rawlings and Hammerman labs for helpful discussion and the study volunteers for their participation. This work was supported by the following funding: L.B.R and L.S (NIH2T32 AI106677), D.J.C. and P.M. (NIH R01AI127726, NIH U19AI125378-S1), J.A.H. (NIH R01AI150178-01S1), H.R.W. (NIH TL1 TR002318), D R. (SCRI, Research Integration Hub Covid-19 Award), M.P. (NIH U01AI142001-02S1; R01AI118803); a Bill & Melinda Gates Foundation grant to N.P.K. (OPP1156262); BWF #1018486 and COVID Pilot grant to M.P.; Emergent Ventures Fast Grant to M.P.

## Author contributions

M.P., L.B.R. and J.N. conceived the study. K.K.T. assisted in cohort recruitment and visit scheduling. J.R., C.S., E.H., L.B.R, J.N. and K.K.T. processed and preserved blood and serum samples. J.N., L.C., and N.P.K. generated proteins and L.B.R. generated and validated tetramer reagents. L.S., J.N. and L.B.R performed ELISAs and L.S. and J.N performed sVNT assays. L.S. analyzed serum data. H.R.W. and J.H. conceived, performed and analyzed innate cell phenotyping experiments. L.B.R. and J.N. performed and analyzed antigen-specific B cell flow cytometry and sorting. K.B.P. and P.M. conceived, performed and analyzed T cell experiments. C.T. sequenced and generated mAb plasmids. Y.C. expressed and purified mAbs. J.E. and E.H. performed PRNT assays. L.B.R, J.N., L.S., K.B.P., P.M. and M.P. drafted the manuscript. All authors helped edit the manuscript. M.P. secured funds and supervised the project.

## Competing Interests

M.P., D.R., J.N., C.T., Y.C. and L.B.R. have filed a patent under the provisional serial no. 63/063,841. Other authors declare no competing interests.

## Additional Information

**Correspondence and requests for materials** should be addressed to M.P.

**Supplementary Information** is available for this paper.

## Extended Data Figure/Table Legends

**Extended Data Table 1.** Study cohort characteristics.

|  | **Previously SARS-CoV-2 Infected (CoV2<sup>+</sup>)** | **Healthy Controls (HC)** | **Historical Negatives (HN)** |
| --- | --- | --- | --- |
| Number of participants<sup>1</sup> | 15 | 17 | 14 |
| Age (years) | 47 (28 – 71) | 42 (24 – 57) | 47.5 (27 – 72) |
| Sex | 27% Male, 73% Female | 47% Male, 53% Female | 21% Male, 79% Female |
| Number of symptoms<sup>2,3</sup> | 5 (1 – 7) | NA | ND |
| Symptom duration (days) | 13 (2 – 31) | NA | ND |
| Time from symptom onset to Visit 1 (days) | 35.5 (19 – 44) | NA | NA |
| Time from symptom onset to Visit 2 (days) | 86 (73 – 110) | NA | NA |
| Time from SARS-CoV-2 positive PCR test to Visit 1 (days) | 28 (20 – 35) | NA | NA |
| Time from SARS-CoV-2 positive PCR test to Visit 2 (days) | 77.5 (64 – 97) | NA | NA |
| Time from Visit 1 to Visit 2 (days) | 46 (39 – 69) | 47 (40 – 61) | NA |
| Year samples drawn | 2020 | 2020 | 2016 – 2019 |

<sup>1</sup> Blood drawn from 14 CoV2<sup>+</sup> and 13 HC at Visit 1 and Visit 2. 1 CoV2<sup>+</sup> and 2 HC were only drawn with Visit 1. 2 HC were only drawn with Visit 2.
<sup>2</sup> All CoV2<sup>+</sup> individuals reported symptoms. 9 HCs reported symptoms and 2 HC had negative SARSCoV-2 PCR results.
<sup>3</sup> The symptoms surveyed were fever, chills, cough, runny nose, fatigue, muscle ache and difficulty breathing.

**Extended Data Table 2.**
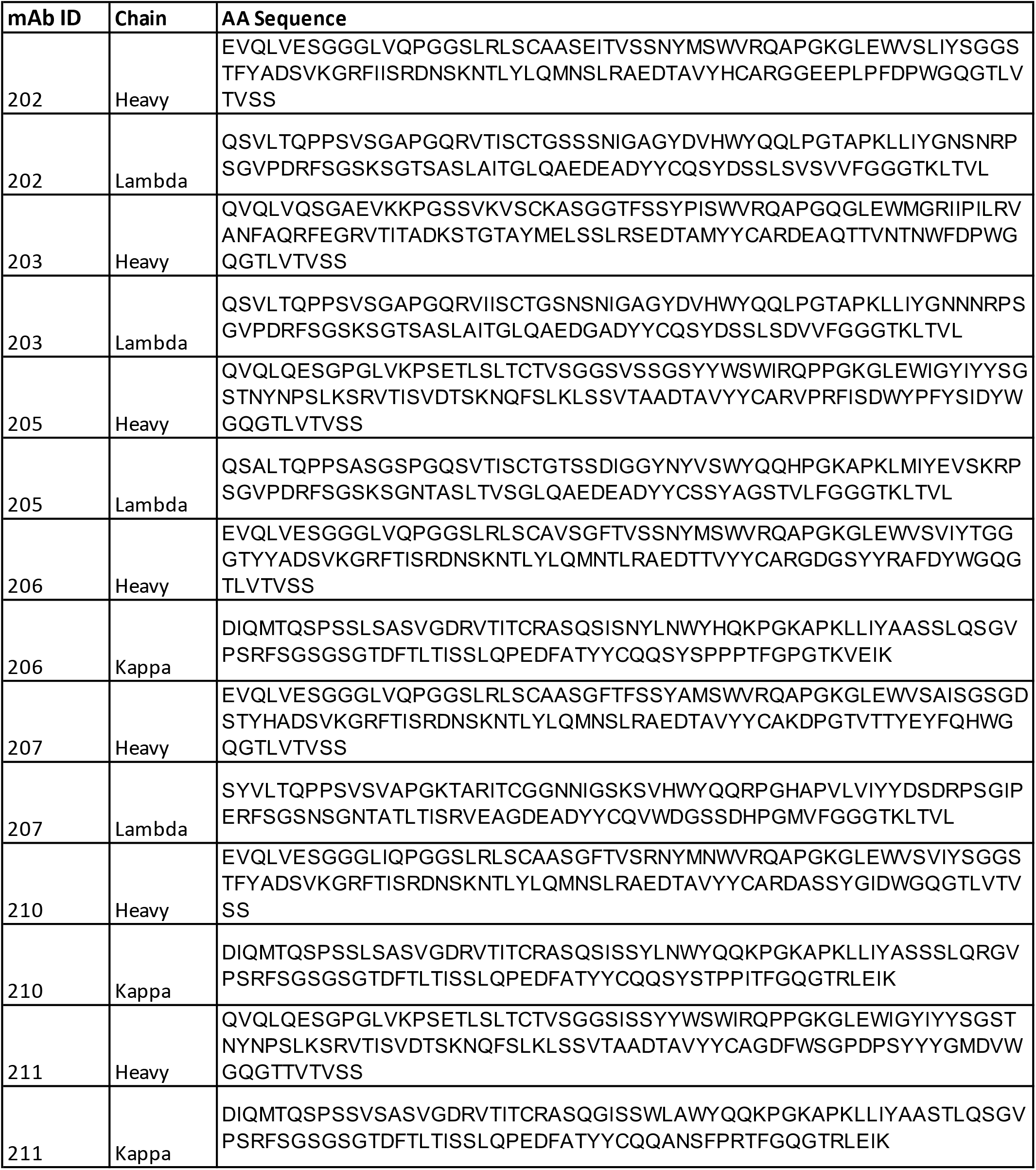
RBD-specific MBC-derived antibody amino acid sequences.

**Extended Data Figure 1.**
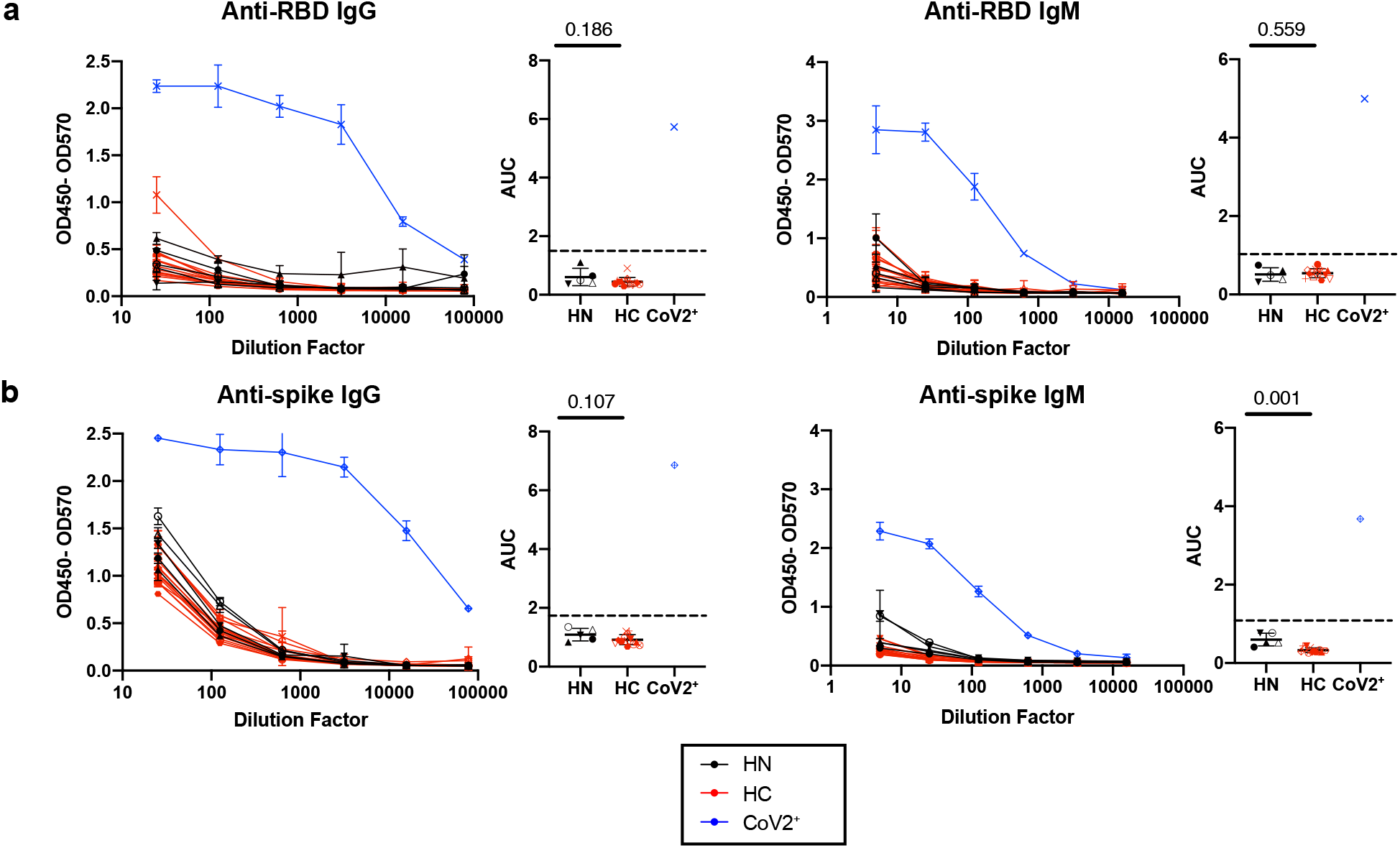
Healthy controls do not have SARS-CoV-2 RBD or spike-specific antibodies. ELISA dilution curves and AUC for anti-RBD **a)** and anti-spike **b)** IgG (left) and IgM (right) in plasma collected prior to the SARS-CoV-2 pandemic (historical negatives, HN, black), during the pandemic (healthy controls, HC, red, at Visit 2), and from individuals previously found PCR positive for SARS-CoV-2 (CoV2^+^, blue, at Visit 1). Dashed line indicates mean + 3 SD of HN AUC values. Statistical significance determined by two-tailed Mann-Whitney tests. Error bars represent mean and SD (HN n=5, HC n=14, IgG CoV2^+^ n=1, IgM CoV2^+^ n=1).

**Extended Data Figure 2.**
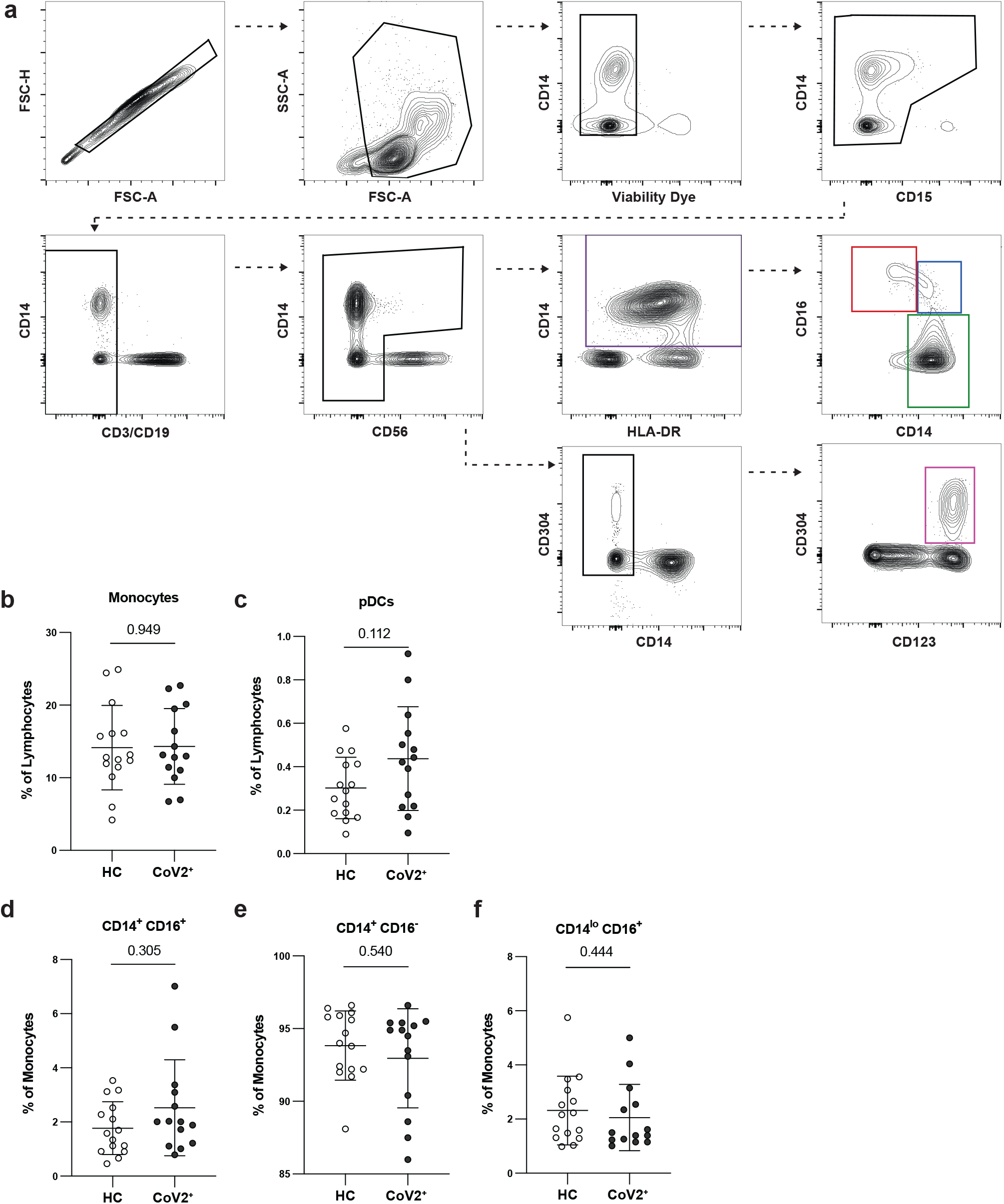
PBMC innate populations in CoV2^+^ and HC individuals are not different at Visit 1. **a)** Flow cytometry gating for CD15^-^CD3^-^CD19^-^CD56^-^HLADR^+^CD14^+^ monocytes (purple gate), which were further divided into CD14^lo^CD16^+^ (red gate), CD14^+^CD16^+^ (blue gate), and CD14^+^CD16^-^ monocytes (green gate), and CD15^-^CD3^-^CD19^-^CD56^-^CD14^-^CD304^+^CD123^+^ plasmacytoid dendritic cells (pDCs) (pink gate). **b)** Percent monocytes and **c)** pDCs of live PBMCs and **d-f**) percent monocyte subsets of monocytes in PBMCs from healthy controls (HC) and previously SARS-CoV-2 infected (CoV2^+^) individuals. Statistical significance determined by twotailed Mann-Whitney tests. Error bars represent mean and SD (HC n=15, CoV2^+^ n=14, 2 experiments).

**Extended Data Figure 3.**
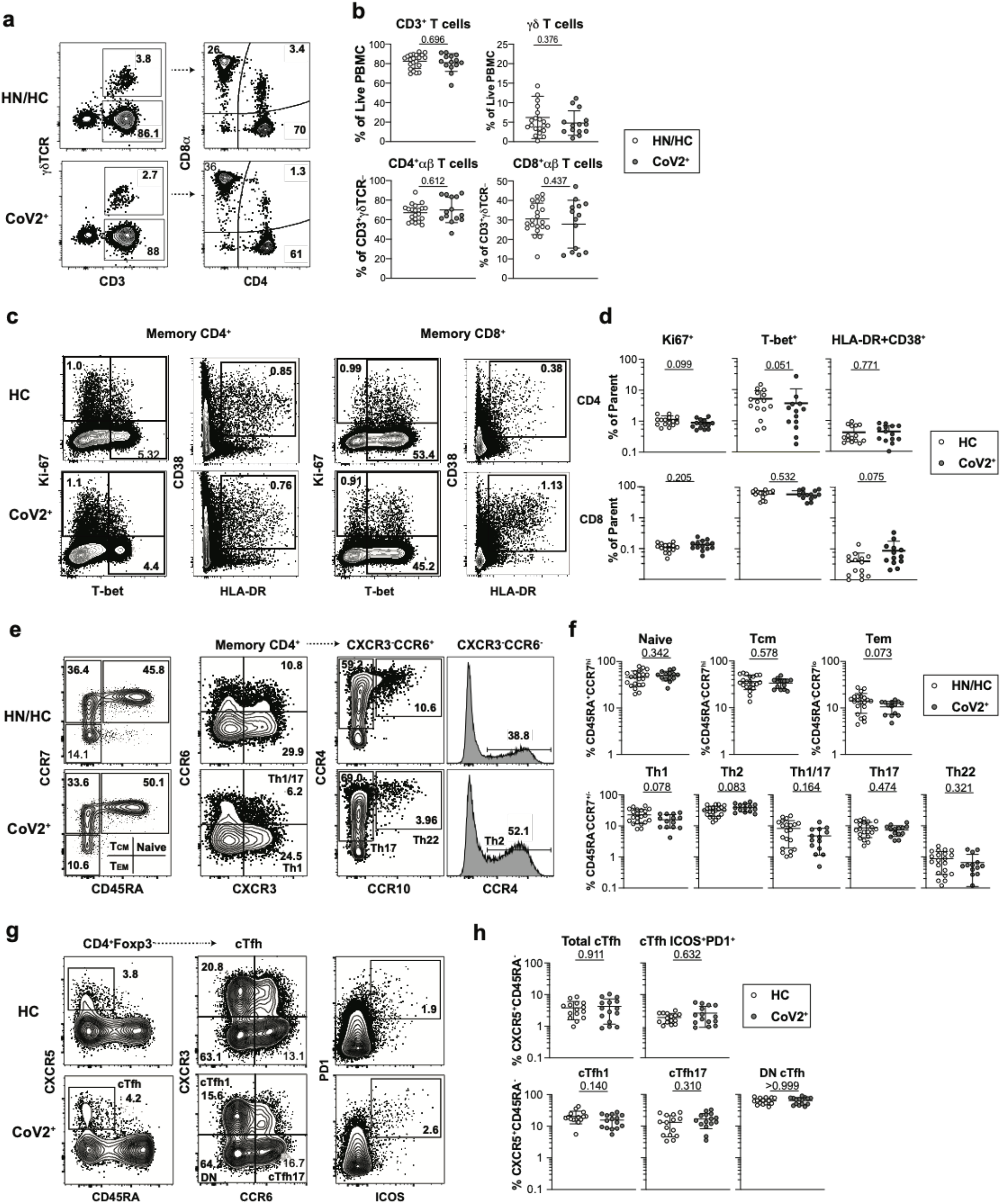
Bulk PBMC T Cells return to immune quiescence by Visit 1. **a)** Representative flow cytometry plots and **b)** frequencies of αβ and γδ T cell subsets at Visit 1 (V1) in PBMCs from historical negative (HN), healthy control (HC) and SARS-CoV-2-recovered (CoV2^+^) PBMCs. **c)** Representative flow cytometry plots and **d)** frequencies of CD4^+^ and CD8^+^ T Cell effector/activation states of total non-naive, memory CD4^+^ or CD8^+^ T Cells (CD45RA^+^CCR7^+/-^) at V1 in HC and CoV2^+^ PBMCs. **e)** Representative flow cytometry plots and **f)** frequencies of CD4^+^ memory and T-helper subsets at V1 in HN/HC and CoV2^+^ PBMCs. **g)** Representative flow cytometry plots and **h)** frequencies of cTfh (CXCR5^+^ CD45RA^-^) and cTfh activation (ICOS^+^PD-1^+^) and helper (CXCR3^+/-^CCR6^+/-^) subsets at V1 in HC and CoV2^+^ PBMCs. Statistical significance determined by two-tailed Mann-Whitney tests. Error bars represent mean and SD (HN n=6, HC n=15, CoV2^+^ n=14, 2 experiments).

**Extended Data Figure 4.**
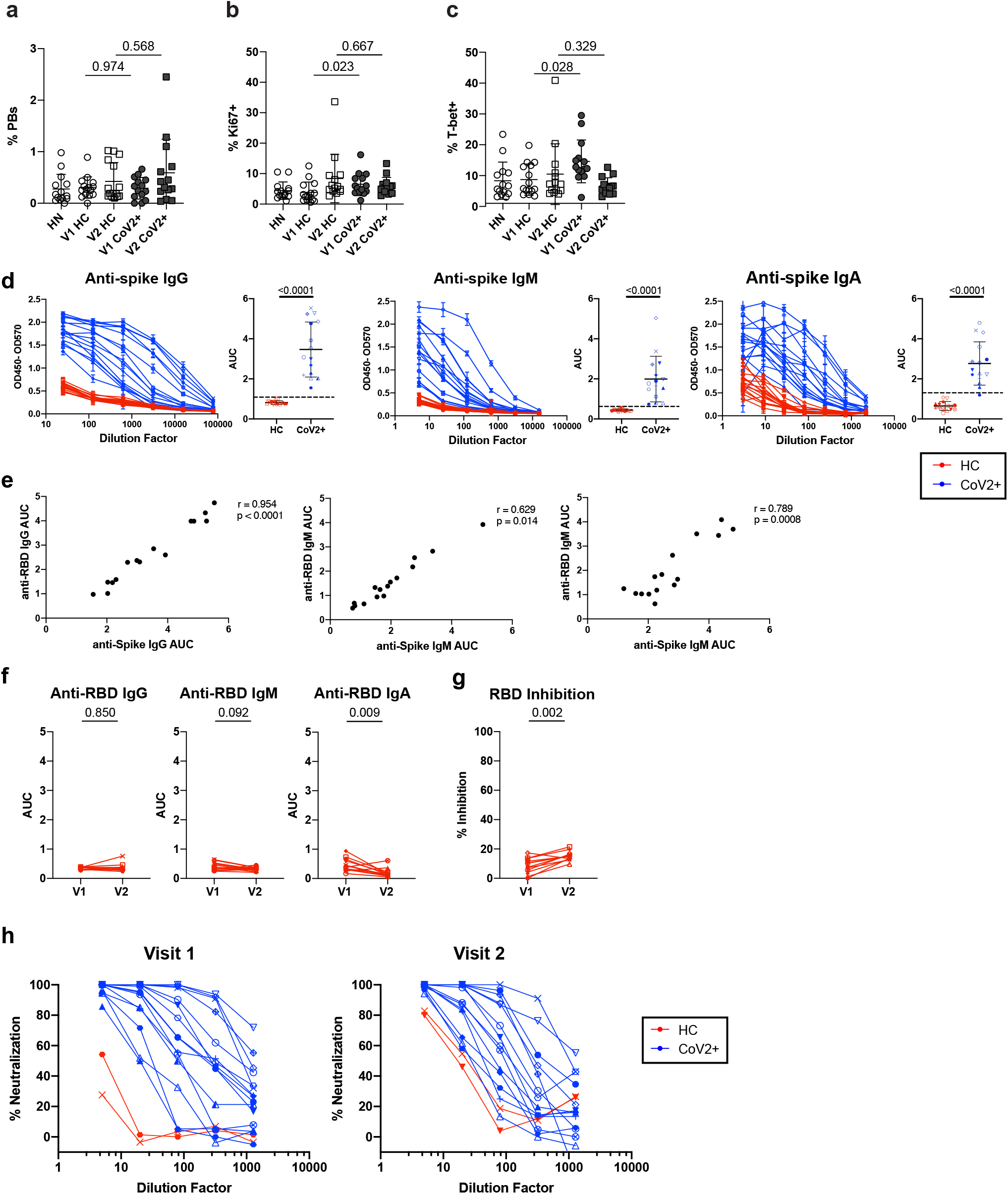
PBMC B Cell and antibody response at two memory time points. **a)** Frequency of plasmablasts (PBs, CD20^-^CD38^hi^) of live, CD3^-^CD14^-^CD16^-^ PBMCs and **b)** cycling cells (Ki67^+^) and **c)** T-bet^+^ cells of live, CD3^-^CD14^-^CD16^-^CD20^+^ PBMCs from healthy control (HC) and SARS-CoV-2-recovered (CoV2^+^) individuals at Visit 1 (V1) and Visit 2 (V2) (HN n=14, V1 HC n=15, V2 HC n=14 (PB) n=15 (Ki67, T-bet), V1 CoV2^+^ n=14, V2 CoV2^+^ n=14, 2 experiments). **d)** ELISA dilution curves and AUC for anti-spike IgG (left), IgM (center), and IgA (right) from HC and CoV2^+^ plasma at V1. Dashed line indicates 3 mean + 3 SD of the HC AUC values. Each symbol is a different individual and is consistent in d), f) and h) (HC n=15, CoV2^+^ n=15). **e)** Spearman correlation of V1 anti-RBD and anti-spike IgG (left), IgM (center), and IgA (right) AUC (HC n=15, CoV2^+^ n=15). **f)** V2 HC AUC values and **g)** RBD inhibition at 1:10 plasma dilution were normalized to V1 CoV2^+^ samples run with V2 samples and value for each HC individual from V1 and V2 are paired (V1 HC n=12, V2 HC n=12). **h)** Percent neutralization dilutions curves determined by PRNT for CoV2^+^ and HC samples (V1 HC n=2, V1 CoV2^+^ n=15, V2 HC n=2, V2 CoV2^+^ n=14). Statistical significance for unpaired data determined by two-tailed Mann-Whitney tests and, for paired data, by two-tailed Wilcoxon signed-rank tests. Error bars represent mean and SD.

**Extended Data Figure 5.**
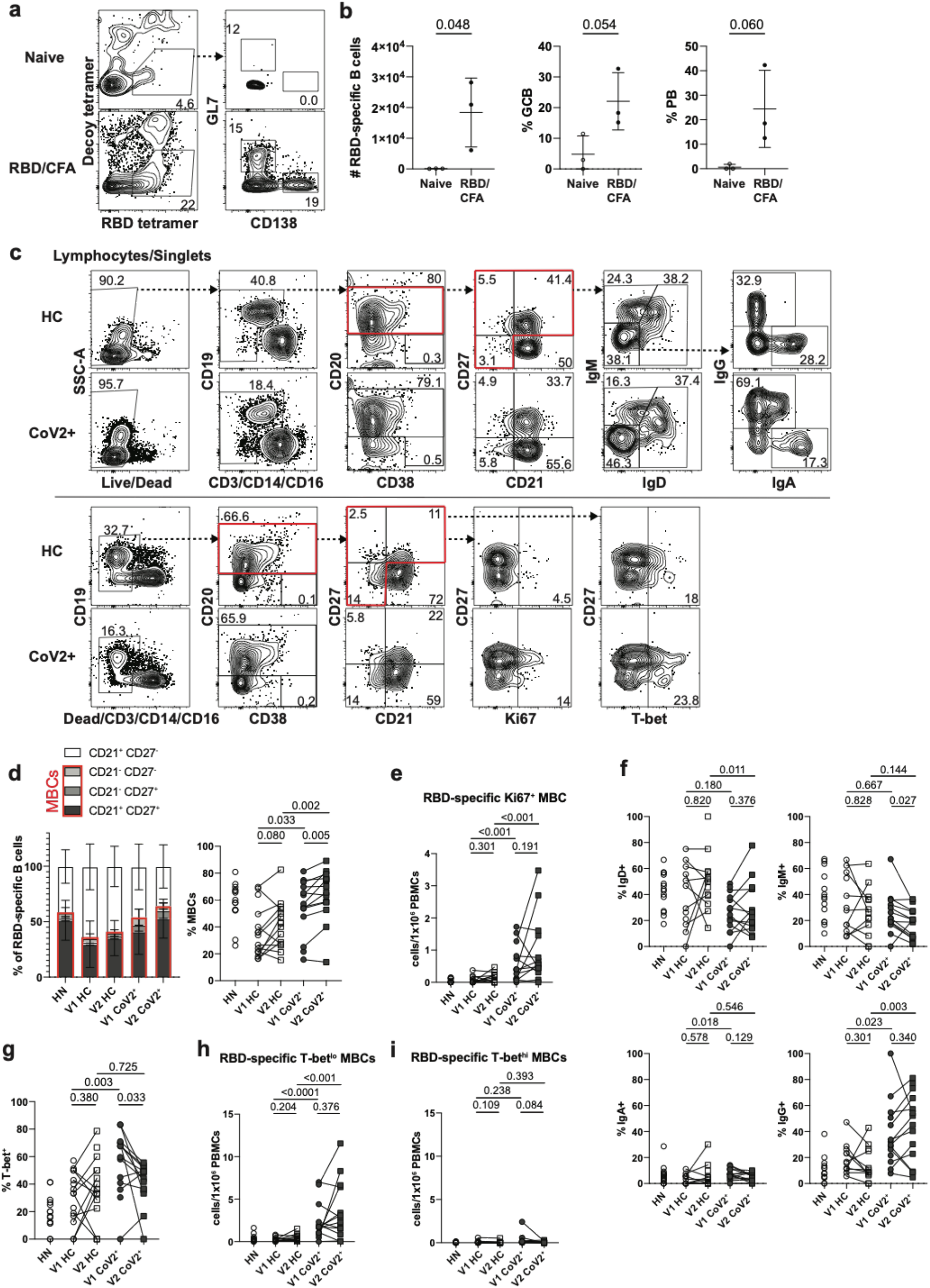
Detecting SARS-CoV-2 RBD-specific B cells in PBMCs. **a,b)** SARS-CoV-2 RBD tetramer detected increased numbers of RBD-specific B cells (Live, CD4^-^ CD8^-^B220^+^and CD138^+^B220^-^RBD tetramer^+^Decoy^-^) which had increased frequencies of germinal center B cells (GCB, GL7+CD138^-^) and plasmablasts (PB, CD138+GL7^-^) in popliteal lymph nodes from mice 7-10 days post-immunization with RBD/CFA compared naive (n=3 mice/treatment in 3 experiments). **c)** Representative flow cytometry gates for phenotyping in Figure 2 set on total B cells from healthy controls (HC) and SARS-CoV-2-recovered (CoV2^+^) in two panels (surface, top; intracellular, bottom). **d)** Left, frequencies of RBD-specific B cells with naive (CD21^+^CD27) and memory (CD21^+^CD27^+^/CD21^-^CD27^+^/CD21^-^CD27^-^) phenotypes from HC and CoV2^+^ PBMCs at Visit 1 (V1) and Visit 2 (V2). Right, frequency of MBCs (populations outlined in red). **e)** Number of cycling (Ki67^+^) RBD-specific MBCs (HN n=14, V1 HC n=12, V2 HC n=13, V1 CoV2^+^ n=15, V2 CoV2^+^ n=14). Frequency of RBD-specific MBCs expressing **f)** indicated BCR isotype (HN n=14, V1 HC n=12, V2 HC n=13, V1 CoV2^+^ n=15, V2 CoV2^+^ n=14) or **g)** T-bet. Number of **h)** resting (T-bet^lo^) and **i)** recently activated (T-bet^hi^) RBD-specific MBCs. Statistical significance for unpaired data determined by two-tailed Mann-Whitney tests and, for paired data, by two-tailed Wilcoxon signed-rank tests. Error bars represent mean and SD (HN n=14, V1 HC n=15, V2 HC n=15, V1 CoV2^+^ n=15, V2 CoV2^+^ n=14, unless otherwise noted, 2 experiments).

**Extended Data Figure 6.**
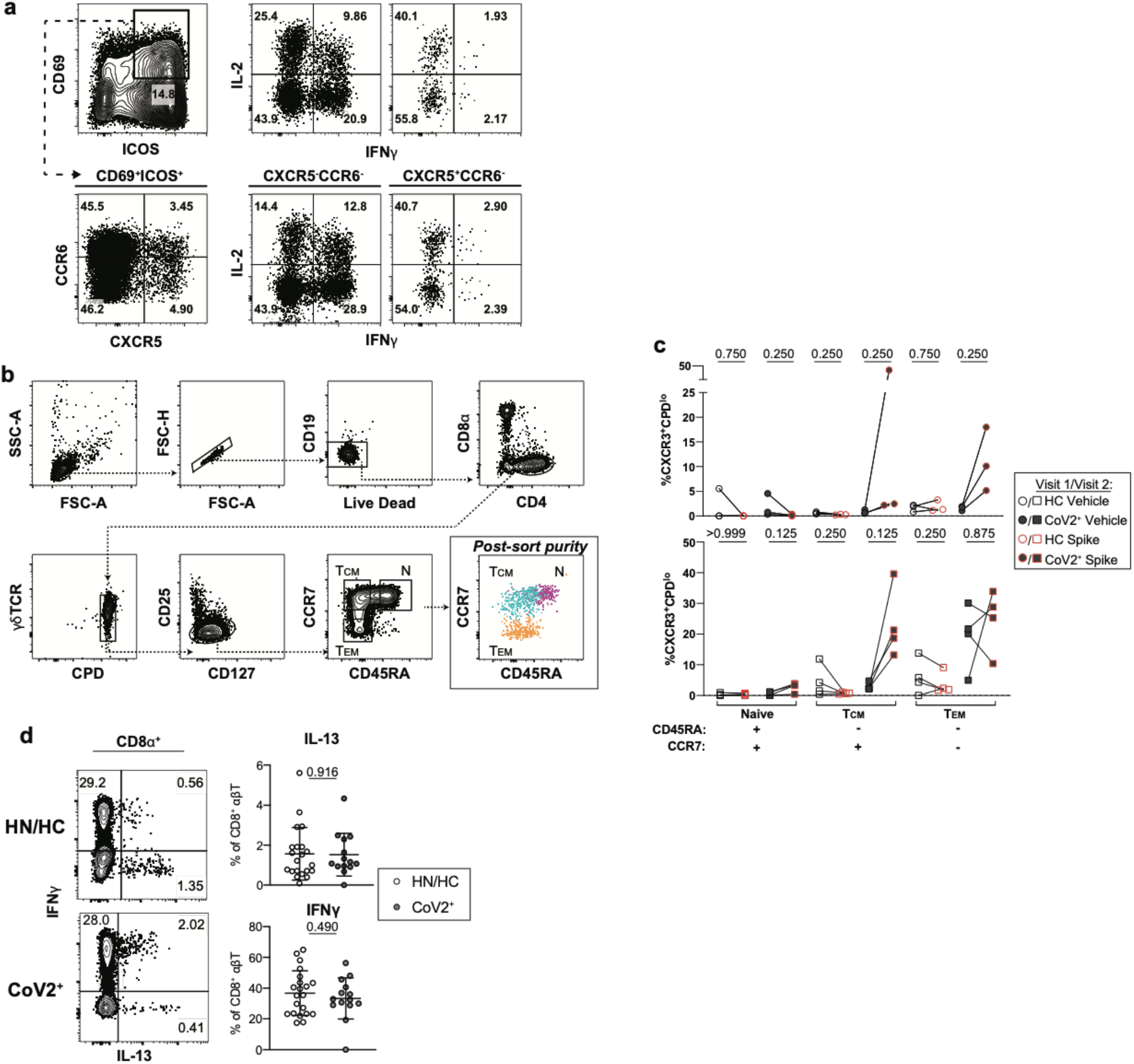
Bulk CD4 and CD8^+^ T cell cytokines expression. **a)** Representative cytokine gating on CD69^+^ICOS^+^ CD4^+^ T Cells from PMA/Ionomycin-activated CD4^+^ cTfh (CXCR5^+^) and non-cTfh (CXCR5^-^) T Cells. **b)** Sorting strategy and **c)** frequency of proliferated (CXCR3^+^CPD^lo^) naive, T central memory (T_CM_), and T effector memory (Tem) cells from HC and CoV2^+^ PBMCs at Visit 1 and Visit 2 after 5-6 days of culture with autologous monocytes and SARS-CoV-2 spike protein or vehicle (V1 HC n=3, V2 HC n=4, V1 CoV2^+^ n=3, V2 CoV2^+^ n=4). **d)** Representative flow cytometry plots of cytokine expression from PMA/Ionomycin-activated CD8^+^ T cells (HN n=6, HC n=15, CoV2^+^ n=14). Statistical significance for unpaired data determined by two-tailed Mann-Whitney tests and, for paired data, by two-tailed Wilcoxon signed-rank tests. Error bars represent mean and SD (2 experiments).

**Extended Data Figure 7.**
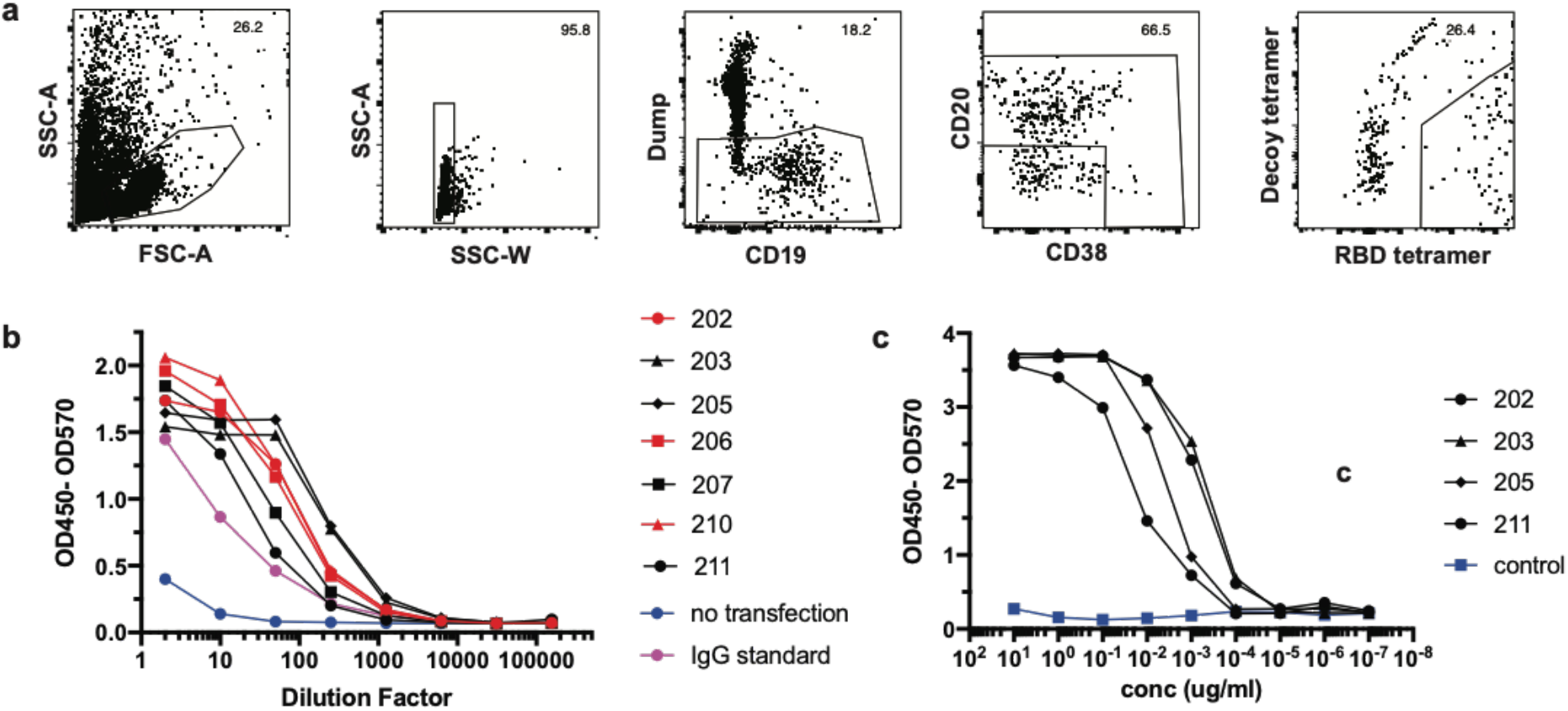
Generation of neutralizing antibodies by RBD-specific MBCs. **a**) Gating strategy for sorting RBD-specific B cells. **b**) IgG ELISA to confirm expression of antibodies in transfected cell culture supernatants. **c**) RBD ELISA of purified monoclonal antibodies. Negative control is an irrelevant *Plasmodium*-specific antibody.

## Notes

### Author Declarations

This study was approved by the University of Washington Institutional Review Board (Gale Lab, IRB 00009810).

### Summary of Updates

Figure 4, E.D. Fig 4 and the corresponding text were revised to accurately report PRNT/IC50 results.

## Main References

1 Wu, Z. & McGoogan, J. M. Characteristics of and Important Lessons From the Coronavirus Disease 2019 (COVID-19) Outbreak in China: Summary of a Report of 72 314 Cases From the Chinese Center for Disease Control and Prevention. JAMA 323, 1239–1242, doi:10.1001/jama.2020.2648 (2020).

2 Ruterbusch, M., Pruner, K. B., Shehata, L. & Pepper, M. In Vivo CD4(+) T Cell Differentiation and Function: Revisiting the Th1/Th2 Paradigm. Annu Rev Immunol 38, 705–725, doi: 10.1146/annurev-immunol-103019-085803 (2020).

3 Knox, J. J., Myles, A. & Cancro, M. P. T-bet(+) memory B cells: Generation, function, and fate. Immunol Rev 288, 149–160, doi: 10.1111/imr.12736 (2019).

4 Kim, C. C., Baccarella, A. M., Bayat, A., Pepper, M. & Fontana, M. F. FCRL5(+) Memory B Cells Exhibit Robust Recall Responses. Cell Rep 27, 1446–1460 e1444, doi:10.1016/j.celrep.2019.04.019 (2019).

5 Nellore, A. et al. Fcrl5 and T-bet define influenza-specific memory B cells that predict long-lived antibody responses. bioRxiv, 643973, doi:10.1101/643973 (2019).

6 Schmidt, M. E. & Varga, S. M. The CD8 T Cell Response to Respiratory Virus Infections. Frontiers in Immunology 9, doi:10.3389/fimmu.2018.00678 (2018).

7 Hoffmann, M. et al. SARS-CoV-2 Cell Entry Depends on ACE2 and TMPRSS2 and Is Blocked by a Clinically Proven Protease Inhibitor. Cell 181, 271–280 e278, doi:10.1016/j.cell.2020.02.052 (2020).

8 Shi, R. et al. A human neutralizing antibody targets the receptor binding site of SARS-CoV-2. Nature, doi:10.1038/s41586-020-2381-y. (2020).

9 Robbiani, D. F. et al. Convergent antibody responses to SARS-CoV-2 in convalescent individuals. Nature 18, 18, doi: https://dx.doi.org/10.1038/s41586-020-2456-9 (2020).

10 Long, Q.-X. et al. Antibody responses to SARS-CoV-2 in patients with COVID-19. Nature Medicine 26, 845–848, doi:10.1038/s41591-020-0897-1 (2020).

11 Wölfel, R. et al. Virological assessment of hospitalized patients with COVID-2019. Nature 581, 465–469, doi: 10.1038/s41586-020-2196-x (2020).

12 Mathew, D. et al. Deep immune profiling of COVID-19 patients reveals distinct immunotypes with therapeutic implications. Science (New York, NY), doi:10.1126/science.abc8511. (2020).

13 Slifka, M. K. & Ahmed, R. Long-term antibody production is sustained by antibody-secreting cells in the bone marrow following acute viral infection. Ann N Y Acad Sci 797, 166–176, doi:10.1111/j.1749-6632.1996.tb52958.x (1996).

14 Knox, J. J. et al. T-bet^+^ B cells are induced by human viral infections and dominate the HIV gp140 response. JCI Insight 2, doi:10.1172/jci.insight.92943 (2017).

15 Ma, H. et al. Serum IgA, IgM, and IgG responses in COVID-19. Cellular & Molecular Immunology 17, 773–775, doi:10.1038/s41423-020-0474-z (2020).

16 Tan, C. W. et al. A SARS-CoV-2 surrogate virus neutralization test based on antibody-mediated blockage of ACE2-spike protein-protein interaction. Nature Biotechnology, doi:10.1038/s41587-020-0631-z (2020).

17 Krishnamurty, A. T. et al. Somatically Hypermutated Plasmodium-Specific IgM(+) Memory B Cells Are Rapid, Plastic, Early Responders upon Malaria Rechallenge. Immunity 45, 402–414, doi:10.1016/j.immuni.2016.06.014 (2016).

18 Ladner, J. T. et al. Epitope-resolved profiling of the SARS-CoV-2 antibody response identifies cross-reactivity with an endemic human CoV. bioRxiv, 2020.2007.2027.222943, doi:10.1101/2020.07.27.222943 (2020).

19 Braun, J. et al. SARS-CoV-2-reactive T cells in healthy donors and patients with COVID-19. Nature, doi:10.1038/s41586-020-2598-9 (2020).

20 Weiskopf, D. et al. Phenotype and kinetics of SARS-CoV-2-specific T cells in COVID-19 patients with acute respiratory distress syndrome. Sci Immunol 5, doi:10.1126/sciimmunol.abd2071 (2020).

21 Knox, J. J., Kaplan, D. E. & Betts, M. R. T-bet-expressing B cells during HIV and HCV infections. Cell Immunol 321, 26–34, doi:10.1016/j.cellimm.2017.04.012 (2017).

22 Weisel, F. & Shlomchik, M. Memory B Cells of Mice and Humans. Annual Review of Immunology 35, 255–284, doi:10.1146/annurev-immunol-041015-055531 (2017).

23 Bentebibel, S. E. et al. Induction of ICOS^+^CXCR3^+^CXCR5^+^ TH cells correlates with antibody responses to influenza vaccination. Sci Transl Med 5, 176ra132, doi:10.1126/scitranslmed.3005191 (2013).

24 Reiss, S. et al. Comparative analysis of activation induced marker (AIM) assays for sensitive identification of antigen-specific CD4 T cells. PLoS One 12, e0186998, doi:10.1371/journal.pone.0186998 (2017).

25 Vinuesa, C. G., Linterman, M. A., Yu, D. & MacLennan, I. C. Follicular Helper T Cells. Annu Rev Immunol 34, 335–368, doi:10.1146/annurev-immunol-041015-055605 (2016).

26 Pepper, M. & Jenkins, M. K. Origins of CD4(+) effector and central memory T cells. Nat Immunol 12, 467–471, doi: 10.1038/ni.2038 (2011).

27 Juno, J. A. et al. Humoral and circulating follicular helper T cell responses in recovered patients with COVID-19. Nature medicine, doi:10.1038/s41591-020-0995-0. (2020).

28 Le Bert, N. et al. SARS-CoV-2-specific T cell immunity in cases of COVID-19 and SARS, and uninfected controls. Nature, doi:10.1038/s41586-020-2550-z (2020).

29 Alsoussi, W. B. et al. A Potently Neutralizing Antibody Protects Mice against SARS-CoV-2 Infection. J Immunol, doi:10.4049/jimmunol.2000583 (2020).

30 Hanke, L. et al. An alpaca nanobody neutralizes SARS-CoV-2 by blocking receptor interaction. bioRxiv, 2020.2006.2002.130161, doi:10.1101/2020.06.02.130161 (2020).

31 Tang, F. et al. Lack of Peripheral Memory B Cell Responses in Recovered Patients with Severe Acute Respiratory Syndrome: A Six-Year Follow-Up Study. The Journal of Immunology 186, 7264–7268, doi:10.4049/jimmunol.0903490 (2011).

32 Wu, L. P. et al. Duration of antibody responses after severe acute respiratory syndrome. Emerg Infect Dis 13, 1562–1564, doi:10.3201/eid1310.070576 (2007).

33 Seow, J. et al. Longitudinal evaluation and decline of antibody responses in SARS-CoV-2 infection. medRxiv, 2020.2007.2009.20148429, doi:10.1101/2020.07.09.20148429 (2020).

34 Perreault, J. et al. Longitudinal analysis of the humoral response to SARS-CoV-2 spike RBD in convalescent plasma donors. bioRxiv, 2020.2007.2016.206847, doi:10.1101/2020.07.16.206847 (2020).

35 Wajnberg, A. et al. SARS-CoV-2 infection induces robust, neutralizing antibody responses that are stable for at least three months. medRxiv, 2020.2007.2014.20151126, doi:10.1101/2020.07.14.20151126 (2020).

36 Isho, B. et al. Evidence for sustained mucosal and systemic antibody responses to SARS-CoV-2 antigens in COVID-19 patients. medRxiv, 2020.2008.2001.20166553, doi:10.1101/2020.08.01.20166553 (2020).

37 Plotkin, S. A. Correlates of protection induced by vaccination. Clin Vaccine Immunol 17, 1055–1065, doi:10.1128/CVI.00131-10 (2010).

38 Grifoni, A. et al. Targets of T Cell Responses to SARS-CoV-2 Coronavirus in Humans with COVID-19 Disease and Unexposed Individuals. Cell 181, 1489–1501 e1415, doi:10.1016/j.cell.2020.05.015 (2020).

39 Rubtsova, K., Rubtsov, A. V., van Dyk, L. F., Kappler, J. W. & Marrack, P. T-box transcription factor T-bet, a key player in a unique type of B-cell activation essential for effective viral clearance. Proc Natl Acad Sci U S A 110, E3216–3224, doi:10.1073/pnas.1312348110 (2013).

40 Lee, Y. K. et al. Late developmental plasticity in the T helper 17 lineage. Immunity 30, 92–107, doi:10.1016/j.immuni.2008.11.005 (2009).

41 Peng, S. L., Szabo, S. J. & Glimcher, L. H. T-bet regulates IgG class switching and pathogenic autoantibody production. Proc Natl Acad Sci U S A 99, 5545–5550, doi:10.1073/pnas.082114899 (2002).

42 Hirota, K. et al. Plasticity of Th17 cells in Peyer’s patches is responsible for the induction of T cell-dependent IgA responses. Nat Immunol 14, 372–379, doi: 10.1038/ni.2552 (2013).

43 Mateus, J. et al. Selective and cross-reactive SARS-CoV-2 T cell epitopes in unexposed humans. Science, doi:10.1126/science.abd3871 (2020).

44 Chandrashekar, A. et al. SARS-CoV-2 infection protects against rechallenge in rhesus macaques. Science, doi:10.1126/science.abc4776 (2020).

45 Erasmus, J. H. et al. Single-dose replicating RNA vaccine induces neutralizing antibodies against SARS-CoV-2 in nonhuman primates. bioRxiv, doi:10.1101/2020.05.28.121640 (2020).

